# The role of social network support in treatment outcomes for medication for opioid use disorder: a systematic review

**DOI:** 10.1101/2020.07.18.20156950

**Authors:** Navin Kumar, William Oles, Benjamin A. Howell, Kamila Janmohamed, Selena T. Lee, Melissa C. Funaro, Patrick G. O’Connor, Marcus Alexander

## Abstract

**Background:** Social connections can lead to contagion of healthy behaviors. Successful treatment of patients with opioid use disorder, as well as recovery of their communities from the opioid epidemic, may lay in rebuilding social networks. Strong social networks of support can reinforce the benefits of medication treatments that are the current standard of care and the most effective tool physicians have to fight the opioid epidemic.

**Methods:** We conducted a systematic review of electronic research databases, specialist journals and grey literature up to August 2020 to identify experimental and observational studies of social network support in patient populations receiving medication for opioid use disorder (MOUD). We place the studies into a conceptual framework of dynamic social networks, examining the role of networks before MOUD treatment is initiated, during the treatment, and in the long-term following the treatment. We analyze the results across three sources of social network support: partner relationships, family, and peer networks. We also consider the impact of negative social connections.

**Results:** Of 5193 articles screened, 46 studies were identified as meeting inclusion criteria (12 were experimental and 34 were observational). 39 studies indicated that social network support, or lack thereof, had a statistically significant relationship with improved MOUD treatment outcomes. We find the strongest support for the positive impact of family and partner relationships when integrated into treatment attempts. We also identify strong evidence for a negative impact of maintaining contacts with the drug-using network on treatment outcomes.

**Conclusions:** Social networks significantly shape effectiveness of opioid use disorder treatments. While negative social ties reinforce addiction, positive social support networks can amplify the benefits of medication treatments. Targeted interventions to reconstruct social networks can be designed as a part of medication treatment with their effects evaluated in improving patients’ odds of recovery from opioid use disorder and reversing the rising trend in opioid deaths.

## Introduction

Opioid dependence continues to be a serious challenge to public and global health. The number of years of life lost due to opioid dependence was 3.6 million worldwide as of 2016 [1]. From 1990 to 2014, opioid prescription sales increased four-fold, and by 2016, 236 million prescriptions were written yearly in the US, enough for every adult American to have one. Recent initiatives to limit over-prescription have mirrored an increase in opioid-related injury and death linked to illicit opioids like heroin and fentanyl [2, 3]. In the 12-month period ending on February 2020, 72,780 opioid deaths were reported, accounting for an 8.4 percent increase from the year before [4]. On average, 199 Americans die every day from opioids.

Increasing access to medication for opioid use disorder (MOUD) treatment programs is a key public health strategy in combating the opioid overdose epidemic [5, 6]. MOUD has shown several benefits such as decreases in mortality, increases in treatment adherence, decreases in heroin use, and augmented health, social and criminal justice outcomes [7, 8, 9]. MOUD refers to several medications, primarily opioid-agonist medications, like methadone and buprenorphine, and opioid-antagonist medications, like naltrexone [10]. Specifically, methadone and buprenorphine block drug-induced euphoria, reduce cravings and binge behaviors, and prevent withdrawal symptoms. Although MOUD approaches are the most efficacious evidence-based treatment for opioid use disorder [11], a significant number of MOUD patients do not have favorable treatment outcomes [12, 13], signaling the need to explore factors beyond medication that might influence treatment. This review fills gaps in the current body of literature on MOUD outcomes in a numbers of areas. A large portion of the research on MOUD has aimed to identify both protective factors predicting successful treatment and risk factors predicting treatment failure. Of this body of work, much of it has focused on either clinical delivery characteristics, like medication dose, or patient-centered descriptors, such as age, comorbid mental health diagnoses, or attitude towards treatment [14, 15, 16]. Novel treatment paradigms, such as open-access models, have gone a long way in decreasing barriers to treatment and improving engagement, but there is still unexplained variability in treatment success and therefore an opportunity for improvement [17]. Less research has focused on how patients’ social environments generally and social networks, in particular, are associated with treatment outcomes.

Phenomena as diverse as cooperation, obesity, drug use, smoking, and alcohol use are associated with social network structure and social tie influence [18, 19, 20, 21]. Similarly, recent work suggests that social networks ties influence patterns of substance use, help-seeking behaviors, and adherence [22, 23, 24]. Previous systematic reviews have detailed psychosocial interventions that complement MOUD [16, 25, 26] and the effectiveness of psychosocial plus pharmacological intervention versus pharmacological intervention alone [27]. However, only a small subset of psychosocial interventions, such as family counseling or network therapy, actively involve patient social relationships in treatment [28]. Overall, there is limited research on the role of social network support on MOUD treatment outcomes, especially on the changes in such networks over the treatment timeline.

This review adopts a social network lens to consider the outcomes of those in MOUD programs. The impact of social ties, and the mosaic of ties which make up social networks, on health has been conceptualized by both social support and social influence [29]. Social support between two individuals has been categorized into different types––for example, emotional support might refer to the amount of love and caring available from another, while instrumental support refers to aid or assistance with tangible needs. High levels of social support are not only important to maintaining relationships but also in maintaining psychological well-being, and it has been shown that the presence or absence of social support is predictive of mental and physical health outcomes [30, 31, 32]. Although this review does not distinguish between social support types, we chose to investigate different types of network relationships with the assumption that they might offer different types of support. Social relationships can also impact individual health through social influence, which can be thought of as the tendency to conform to the attitudes and behaviors of those with whom you are connected in a network [29]. Recent work to investigate how individual drug-use is influenced by social contacts has found that there exists strong peer-to-peer influence in initiation and continuation of drug-use [22, 33, 34, 35]. While this suggests that outcomes might be improved if those in treatment have little contact with other substance users, the influence of social ties may be more complicated; positive relationships with others may buffer the effects of stress, for example, but these relationships may be with other substance users, who can model drug-use or provide cues for relapse.

Given the proposed influence of social network ties on treatment, the selected studies were classified by relationship type investigated––family, partner, or peer––as well as the time in the treatment timeline at which social support information was elicited––pre-treatment, during treatment, or post-treatment. With both social support and social influence in mind, studies were also classified as assessing positive (e.g., presence of emotional support, abstinence-positive social influence) and negative impacts of social ties (e.g., lack of support or high conflict, drug-use-encouraging social influence) among those in treatment. The purpose of this paper is to review existing scientific evidence on the following research question: For MOUD patients, what influence does social network support, or lack thereof, have on MOUD treatment outcomes? This review seeks to provide policymakers, administrators, practitioners and researchers with a systematic and reproducible strategy to query the literature around the role of social network support on MOUD treatment outcomes.

## Methods

We conformed to frameworks and standard tools of the Preferred Reporting Items for Systematic Review and Meta-analysis Protocols (PRISMA-P) [36, 37, 38] and Synthesis without meta-analysis (SWiM) guidelines [39]. The protocol was pre-registered on PROSPERO (CRD42018095645) on May 24, 2018.

### Search strategy

A systematic search of the literature was performed on August 30, 2018 and updated on August 3, 2020 to capture any new studies. Databases searched included Ovid MEDLINE, Embase, APA Psycinfo, and Sociological Abstracts. An experienced medical librarian (MF) consulted on methodology and ran a medical subject heading (MeSH) analysis of known key articles provided by the research team [mesh.med.yale.edu]. In each database, scoping searches were used alongside an iterative process to translate and refine the searches. To maximize sensitivity, the formal search used a minimal controlled vocabulary terms and synonymous free-text words to capture concepts for “social network support” and “medication for opioid use disorder” (see **??** for full list of search terms). The search was limited to English language. No date limit was applied. In addition, the authors searched references in previous reviews/guidelines, and clinicaltrials.gov.

#### Inclusion/exclusion criteria

Studies that met the following criteria were included:

- randomized controlled trials, quasi-experimental studies and observational studies published in peer-reviewed journals; other scientific publications (e.g., scientific monographs); non-peer reviewed journals and grey literature (technical reports, conference papers). Studies excluded from review were case reports, systematic literature reviews, qualitative studies, opinion pieces, editorials, comments, news articles, and letters.
- participants sought treatment for opioid use or met criteria for opioid abuse, opioid dependence or opioid use disorder.
- one or more variants of MOUD were offered (e.g., methadone, buprenorphine, naltrexone).
- the study reported social network support (e.g., family/partner/peer support, social network interventions [31, 32]) as interventions or as predictors for the outcome.
- the study reported a form of adherence to MOUD as an outcome (e.g., medication adherence, concurrent abstinence during treatment, program retention).
- the study did not exclusively look at peer support groups (e.g., Narcotics Anonymous) or group psychotherapy as a form of social support. Studies involving peer support group attendance as a form of treatment are both numerous and valuable and are therefore significant enough to be deserving of their own review.

### Category assignment

Studies were assigned into the following categories based on when social network support was assessed: pre-treatment, during treatment, and post-treatment. Social network support was assessed at multiple time points in some studies. Pre-treatment indicated studies where social network support was assessed at baseline, before patients commenced an MOUD program. During treatment connoted studies where social network support was assessed while participants were actively participating in an MOUD program. Post-treatment denoted studies where social network support was determined after patients had completed the prescribed MOUD program. The intention of this categorization was to investigate whether the presence or lack of social support seems to be more important at a certain point in the treatment timeline and, more generally, whether there are gaps in the literature at particular points of the timeline.

Studies were assigned to one of the following social network support categories: family, partner, peer. Multiple forms of social network support were assessed in some studies. Family social network support connoted studies where family members such as parents, siblings, or children provided support to the patient. Partner social network support denoted studies where partners, married or otherwise, provided support to the patient. Peer social network support denoted studies where peers such as friends, colleagues or other patients provided support. Studies which solely assessed the impact of mutual peer support groups (e.g., Narcotics Anonymous, Alcoholics Anonymous) or group psychotherapy were excluded. The authors acknowledge the importance of such programs and the wealth of literature on their potential efficacy, and therefore believe that they warrant their own review. It should be noted that some studies describe general family relationships, which may be inherently inclusive of a partner; therefore, any studies which noted general family relationships were assumed to assess both family and partner relationships in our category assignment. Studies were also assigned as assessing either negative or positive social network support. Some studies assessed both positive and negative social network relationships. Examples of positive social network ties include those that provide the knowledge that one is cared for or that encourage treatment, such as family members who sponsor a patient through an MOUD program. Examples of negative social network ties include those that may be sources of conflict or that encourage drug-use, such as individuals going through marital status changes or drug-using peers.

Category assignment choices are driven by our hypothesized narrative for the role of social network structure in MOUD outcomes based on previous literature, shown in Fig 1. In this narrative, successful treatment is accompanied by a transition in social network ties from pretreatment to post-treatment away from the drug-using network and towards the non-using support network, typically formed by close family and friends, including members of a sober network that may be part of a recovery program. The idea that treatment outcomes are related to social relationships, which either reinforce drug use or reinforce abstinence, is long-standing and provides a rationale for the methods of many of the studies under review here [40, 41, 42, 43, 44]. However, no other study has systematically applied such a narrative of social network change over the treatment timeline to frame outcomes reported in the literature.

**Figure 1:**
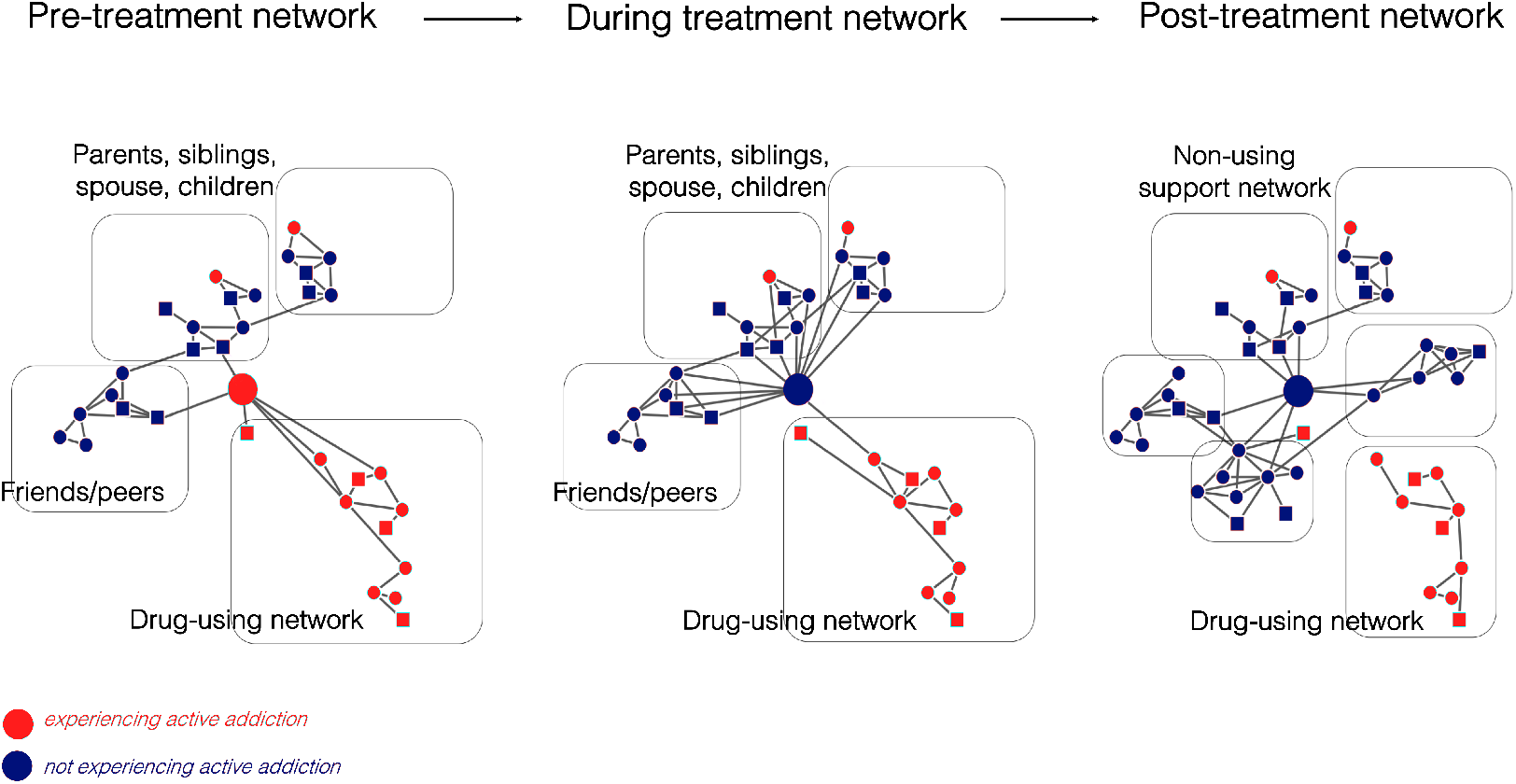
Social network change over treatment timeline. Narrative framework for how dynamic social network changes might accompany successful MOUD treatment outcomes, before, during, and after treatment, with particular emphasis on developing more non-substance-using relationships

### Outcomes

Examples of primary outcomes were 1) MOUD retention which refers to time in treatment or length of stay; 2) MOUD adherence which pertains to medication (days the patient took their MOUD etc.); 3) opioid or other illicit drug use (cocaine, methamphetamine etc.), defined as the percentage of urine samples negative for opioids and/or self-reported drug use.

### Data extraction, review methods, quality assessments and data synthesis

A standardized template was utilized to extract data from each study. More detail on data extraction, review methods, quality assessments and data synthesis can be found in **??**.

## Results

### Included studies

Results from the study selection process are indicated in Fig 2 and general study characteristics are displayed in Tables 1 and 2. Systematic searches yielded 7995 papers imported for screening, with 5193 studies screened for review (2802 duplicates, see Fig 2). Screening yielded 225 articles for full-text review by two independent reviewers. Fourty-six studies were deemed relevant to the review, summarized in Table 1. Twelve were experimental studies, 34 were observational studies. Thirty-nine studies indicated that at least one variant of social network support, or lack thereof, assessed in the study had a statistically significant relationship with MOUD treatment outcomes. Twenty-four studies assessed social network support pre-treatment, 25 during treatment, and five post-treatment. Forty-eight studies detailed family social network support, 46 indicated partner support, and 22 explored peer social network support. The United States (19 studies) and China (10 studies) were the most represented nations. Treatment and comparison groups were all drawn from opioid-dependent populations. Thirty-six interventions involved maintenance on methadone, 4 involved naltrexone, 2 involved methadone or buprenorphine, 1 involved methadone or buprenorphine/naloxone, 1 involved methadone or LAAM (levo-alpha-acetyl-methadol), and 2 involved methadone or naltrexone.

**Table 1:** Study characteristics related to design of study, MOUD drug used, target population, outcome, number of participants, study length

| Author, date | Place | Study Design | MOUD drug | Population | Outcomes | Sample characteristics (N, % male, age, ethnicity) | Study length |
| --- | --- | --- | --- | --- | --- | --- | --- |
| Anton et al., 1981 | USA | Observational, retrospective study | Naltrexone | Opioid dependent self-referred patients | Naltrexone maintenance for three months or more | N: 65 (total),<br>40 (naltrexone + HIP),<br>25 (naltrexone + HIP + MFT)<br>% male: 89<br>Mean age: 26.1<br>Ethnicity: 54% White; 46% Black | 2 years |
| Carroll et al., 2001 | USA | RCT | Naltrexone | Opioid dependent patients who had completed outpatient detoxification | Compliance with naltrexone treatment | N: 127 (total),<br>44 (naltrexone),<br>35 (naltrexone + CM),<br>48 (naltrexone + CM + SO)<br>% male: 76<br>Mean age: 32<br>Ethnicity: 77% White | Unclear |
| Catalano et al., 1997 | USA | RCT | Methadone | Opioid dependent parents who had been in methadone treatment for a minimum of 90 days and have one or more children between the ages of 3 and 14 years old living with them at least 50% of the week | Parent self-reported drug-use | N: 144 (total),<br>62 (control),<br>82 (FOF)<br>% male: 25<br>Mean age: 35.4<br>Ethnicity: 77% White; 18% Black; Mixed/Other: 5% | 3 years |
| Catalano et al., 1999 | USA | RCT | Methadone | Opioid dependent parents who had been in methadone treatment for a minimum of 90 days and have one or more children between the ages of 3 and 14 years old living with them at least 50% of the week | Self-reported frequency of use of marijuana, cocaine, opiates and drug use in previous month | N: 144 (total),<br>62 (control),<br>82 (FOF)<br>% male: 25<br>Mean age: 35.4<br>Ethnicity: 77% White; 18% Black; Mixed/Other: 5% | 3 years |
| Cerovecki et al., 2013 | Croatia | Observational, prospective study | Methadone | Opioid dependent patients treated in family medicine setting | Fatal outcome risk factors like quality or relationships and continuation of drug use during previous therapeutic attempts | N: 287<br>% male: NA<br>Mean age: NA<br>Ethnicity: NA | 12 years |

Table 1: Study Characteristics (continued)
| Author, date | Place | Study Design | MOUD drug | Population | Outcomes | Sample characteristics (N, % male, age, ethnicity) | Study length |
| --- | --- | --- | --- | --- | --- | --- | --- |
| Davila Torres, 2011 | USA | Observational, retrospective study | Methadone | Latino opioid dependent patients | Retention and treatment program completion | N: 291<br>% male: 67<br>Mean age: NA<br>Ethnicity: 100% Hispanic | 1 year |
| Day et al., 2013 | UK | Cross-sectional analysis | Methadone or buprenorphine | Opioid dependents | Heroin use in the previous month | N: 118<br>% male: 79<br>Mean age: 35.5<br>Ethnicity: 73% White; 14% South Asian; 4% Black, 8% Mixed | 6 months |
| Day et al., 2018 | UK | RCT | Methadone or buprenorphine | Opioid dependent individuals who had been prescribed methadone or buprenorphine for the past 12 months and had reported heroin use on one or more days 28 days prior to beginning of the study | Number of days of heroin use in the past month | N: 83 (total), 30 (TAU), 26 (TAU + B-SNBT), 27 (TAU + PGS)<br>% male: 79<br>Mean age: 35.5<br>Ethnicity: 73% White; 14% South Asian; 4% Black; 8% Mixed | 1 year |
| Fals-Stewart et al., 2001 | USA | RCT | Methadone | Opioid dependent male partners who had been married for at least 1 year or in a stable common-law relationship for at least 2 years who had to refrain from seeking additional substance abuse treatment except for self-help meetings like Alcoholics Anonymous | Urine tests | N: 43 (total), 22 (individual counseling), 21 (BCT)<br>% male: 100<br>Mean age: 38.1<br>Ethnicity: 18% White; 15% Black; 3% Hispanic | 3 years |

Table 1: Study Characteristics (continued)
| Author, date | Place | Study Design | MOUD drug | Population | Outcomes | Sample characteristics (N, % male, age, ethnicity) | Study length |
| --- | --- | --- | --- | --- | --- | --- | --- |
| Fals-Stewart and O'Farrell, 2003 | USA | RCT | Naltrexone | Opioid dependent individuals living with at least one parent/spouse/intimate partner/family member willing to participate and did not have a current substance use disorder or meet DSM-III-R criteria for schizophrenia, bipolar disorder, or psychosis | Percentage of opioid-free urines | N: 124 (total), 62 (individual counseling), 62 (BFC)<br>% male: 100<br>Mean age: 32.9<br>Ethnicity: 66% White; 26% Black; 3% Hispanic; 5% Other | 1 years |
| Feng et al., 2018 | China | Observational, retrospective study | Methadone | Opioid dependent patients enrolled in methadone clinic | Self-reported heroin use or a positive urine morphine test result | N: 2446<br>% male: 79<br>Mean age: NA<br>Ethnicity: NA | 1 year |
| Goehl et al., 1993 | USA | Observational, prospective study | Methadone | Adult patients in a community-based, university-affiliated methadone maintenance treatment program in New York | Abstinence (percentage of positive urinalysis in 3 month period) | N: 70<br>% male: 67<br>Mean age: 35<br>Ethnicity: 64% White/Caucasian, 17% African-American, 19% Hispanic-American | 3 months |
| Gogineni et al., 2001 | USA | Observational, prospective study | Methadone | Opioid dependent patients enrolled in MMT clinic | Continued opioid use | N: 252<br>% male: 56<br>Mean age: 40.3<br>Ethnicity: 64% White; 22% Black; 9% Latino/Hispanic; 2% Native American; 3% Other | Unclear |
| Grey et al., 1986 | USA | Observational, prospective study | Methadone or naltrexone | Opioid dependent patients who had been opiate-free for 7 days prior to treatment (for naltrexone) and opiate dependents for at least 1 year for methadone maintenance treatment | Retention (number of required clinic appointments kept) | N: 60<br>% male: 73<br>Mean age: NA<br>Ethnicity: 58.3% White; 11.7% Black; 30.0% Hispanic | 3 months |

Table 1: Study Characteristics (continued)
| Author, date | Place | Study Design | MOUD drug | Population | Outcomes | Sample characteristics (N, % male, age, ethnicity) | Study length |
| --- | --- | --- | --- | --- | --- | --- | --- |
| Griffith et al., 1998 | USA | Observational, retrospective study | Methadone | Opioid drug users who were admitted to three publicly funded methadone treatment clinics in Texas participating in the Drug Abuse Treatment for AIDS-Risk Reduction (DATAR) project from April 1990 through August 1993 | Deviance at 1-year follow-up (Three measures were of opioid use and included self-reported heroin use, self-reported speedball use, and the urinalysis results) | N: 960<br>% male: 69<br>Mean age: 37<br>Ethnicity: 37% White; 21% Black; 37% Hispanic; 2% Other | 3 years |
| Gu et al., 2013 | China | RCT | Methadone | Heroin dependent patients recently admitted to the three participating MMT clinics | Attrition from the MMT service, which was defined as a failure to visit the MMT clinic consecutively for at least 1 month immediately prior to the study's completion date | N: 288 (total),<br>146 (control),<br>142 (psychosocial intervention)<br>% male: 92<br>Mean age: NA<br>Ethnicity: 98.9% Han | 1 year |
| Gu et al., 2014 | China | Case-crossover design | Methadone | Opioid dependent patients | Methadone maintenance clinic nonattendance | N: 131<br>% male: 88<br>Mean age: NA<br>Ethnicity: NA | 1 month |
| Heinz et al., 2009 | USA | Observational, retrospective study | Methadone | Opioid dependent patients with a history of cocaine use | Positive urinalysis tests for cocaine and opiates | N: 635<br>% male: 54<br>Mean age: 39.2<br>Ethnicity: 39.5% White; 59% Black; 1.5% Other | 6 years |
| Hikmayani et al., 2012 | Indonesia | Observational, prospective study | Methadone | Heroin dependent patients | Retention rate in methadone management treatment clinics | N: 98<br>% male: NA<br>Mean age: 31.6<br>Ethnicity: NA | 1 year and 6 months |
| Hoang et al., 2015 | Vietnam | Observational, prospective study | Methadone | Opioid dependent patients | Concurrent heroin use after methadone treatment initiation | N: 965<br>% male: 95<br>Mean age: 32.3<br>Ethnicity: NA | 10 months |

Table 1: Study Characteristics (continued)
| Author, date | Place | Study Design | MOUD drug | Population | Outcomes | Sample characteristics (N, % male, age, ethnicity) | Study length |
| --- | --- | --- | --- | --- | --- | --- | --- |
| Hoang et al., 2018 | Vietnam | Observational, retrospective study | Methadone | Opioid dependent patients | Concurrent heroin use after methadone treatment initiation | N: 500<br>% male: 97<br>Mean age: NA<br>Ethnicity: NA | 5 years |
| Hojjat et al., 2017 | Iran | RCT | Methadone | Individuals with a substance dependent husband with no history of drug dependence | Relapse rate | Total N: 50 (total), 25 (control), 25 (partner education intervention)<br>% male: 100<br>Mean age: 31.5<br>Ethnicity: NA | 1 year and 2 months |
| Kidorf, 2018 | Unclear | RCT, single group design | Methadone | Opioid dependent individuals recruited from a community syringe exchange or from a methadone maintenance program | Rate of alcohol and other drug use | N: 66<br>% male: 47<br>Mean age: 43.2<br>Ethnicity: NA | Unclear |
| Lee et al., 2015 | Taiwan | Observational, prospective study | Methadone | Heroin dependent patients enrolled in MMT clinic | Treatment retention | N: 177<br>% male: 60<br>Mean age: 37.2<br>Ethnicity: NA | Unclear |
| Li et al., 2012 | China | Observational, prospective study | Methadone | Patients enrolled in a participating MMT clinic | Concurrent heroin use during MMT | N: 178<br>% Male: 66<br>Mean age: NA<br>Ethnicity: NA | Unclear |
| Lin et al., 2011 | China | Cross-sectional study | Methadone | Opioid dependent patients enrolled in MMT clinic | Self-reported drug use and urine tests | N: 560<br>% male: 84<br>Mean age: 33<br>Ethnicity: NA | 7 months |
| Lin et al., 2013 | Taiwan | Observational, prospective study | Methadone | Opioid dependent patients enrolled in MMT clinic | Retention rate in MMT clinics | N: 368<br>% male: 86<br>Mean age: 37.2<br>Ethnicity: NA | 1 year |

Table 1: Study Characteristics (continued)
| Author, date | Place | Study Design | MOUD drug | Population | Outcomes | Sample characteristics (N, % male, age, ethnicity) | Study length |
| --- | --- | --- | --- | --- | --- | --- | --- |
| Lister et al., 2017 | USA | Observational, prospective study | Methadone | African American individuals with past year opioid dependence, 30-day opioid use prior to enrollment in MMT, and positive urinalyses at intake | Number of days retained in treatment; Proportion of positive opioid UDS during first month of treatment | N: 101<br>% Male: 0<br>Mean age: 50<br>Ethnicity: 100% African American | 1 year and 3 months |
| Lundgren et al., 2007 | USA | Observational, prospective study | Methadone | Opioid dependent patients enrolled in MMT clinic | Longest consecutive stay in MMT clinic | N: 8258<br>% male: 66<br>Mean age: 36.5<br>Ethnicity: 68.6% White; 25.9% Latino; 5.5% Black | 6 years |
| Mutasa, 2001 | UK | Observational, prospective study | Methadone | Opioid dependent individuals living within catchment area | MST noncompliance (misuse drugs) | N: 45<br>% male: 67<br>Mean age: NA<br>Ethnicity: 88.89% White; 6.67% South Asian; 4.44% Black | Unclear |
| Nguyen et al., 2017 | Vietnam | Observational, prospective study | Methadone | Opioid dependent patients enrolled in MMT clinic | Treatment adherence | N: 241<br>% male: NA<br>Mean age: NA<br>Ethnicity: NA | 4 months |
| Roozen et al., 2003 | Netherlands | RCT | Methadone or naltrexone | Opioid dependent patients | Abstinence for a 6-month treatment period | N: 44 (total), 24 (naltrexone), 20 (methadone)<br>% male: 84.7<br>Mean age: 30.2<br>Ethnicity: NA | 2 years |
| Rothenberg et al., 2002 | US | Prospective study | Naltrexone | Opioid dependent individuals with a significant other who could commit to participating in treatment | Retention in treatment | N: 82<br>% male: 77<br>Mean age: 33.6<br>Ethnicity: 64% White; 25% Hispanic; 11% Black | Unclear |

Table 1: Study Characteristics (continued)
| Author, date | Place | Study Design | MOUD drug | Population | Outcomes | Sample characteristics (N, % male, age, ethnicity) | Study length |
| --- | --- | --- | --- | --- | --- | --- | --- |
| Sarasvita et al., 2012 | Indonesia | Observational, prospective study | Methadone | Opioid dependent patients enrolled in MMT clinic | Duration of treatment in days | N: 178<br>% male: 90<br>Mean age: 27.2<br>Ethnicity: 32.6% Javanese; 16.3% Sudanese; 4.5% Bataknese | 1 year and 6 months |
| Shen et al., 2018 | China | Observational, prospective study | Methadone | Opioid dependent patients enrolled in MMT clinic | Positive urinalysis tests for heroin use | N: 324<br>% male: 77<br>Mean age: 45.2<br>Ethnicity: 83% Han | 1 year |
| Skinner et al., 2011 | USA | Observational, prospective study | Methadone | Opioid addicted adults with 3- 14-year old children enrolled in MMT | 30-day drug use | N: 144<br>% Male: 26<br>Mean age: 48.5<br>Ethnicity: 78% White; 17% Black; 2% Other; 2% Mixed | 12 years |
| Smith, 2002 | USA | Observational, prospective study | Methadone | Male veteran literate opioid dependents individuals | Retention as measured by length of time in treatment | N: 80<br>% male: 60<br>Ethnicity: 60% Black; 38.8% White; 1.2% Latino | 12 weeks |
| Tang, 2016 | China | Observational, prospective study | Methadone | Opioid dependent patients with a history of quitting attempts | Treatment adherence | N: 523<br>% male: 76<br>Mean age: 38.5<br>Ethnicity: NA | 9 months |
| Torrens et al., 1996 | Spain | Observational, prospective study | Methadone | Opioid dependent patients | Retention rate | N: 370<br>% male: 66<br>Mean age: 29.6<br>Ethnicity: NA | 2 years and 10 months |
| Tran et al., 2018 | Vietnam | Observational, prospective study | Methadone | Opioid dependent patients enrolled in MMT clinic | Self-reported medication adherence | N: 510<br>% male: 98<br>Mean age: 36.6<br>Ethnicity: NA | 5 months |
| Wasserman et al., 2001 | USA | Observational, prospective study | Methadone or LAAM | Opioid dependent patients with at least 1 month enrollment in methadone or LAAM maintenance | Biochemically confirmed opiate and cocaine abstinence | N: 128<br>% male: 55<br>Mean age: 45<br>Ethnicity: 50% White | Unclear |

Table 1: Study Characteristics (continued)
| Author, date | Place | Study Design | MOUD drug | Population | Outcomes | Sample characteristics (N, % male, age, ethnicity) | Study length |
| --- | --- | --- | --- | --- | --- | --- | --- |
| Yandoli et al., 2002 | UK | RCT | Methadone | Patients with at least six months' duration of opioid dependence who agreed to be seen with their partner/family during treatment if required | Frequency of opiate use (allocated as opiate free, occasional users, regular users) | N: 119 (total), 40 (low-contact), 38 (standard), 41 (family counseling)<br>% male: 63<br>Mean age: 28.2<br>Ethnicity: NA | 14 months |
| Yang et al., 2013 | China | Observational, retrospective study | Methadone | Opioid dependent patients enrolled in MMT clinic | Retention rate in MMT clinic | N: 2728<br>% male: 73<br>Mean age: 36.4<br>Ethnicity: NA | 4 years and 9 months |
| Zhou et al., 2017 (1) | China | Observational, ambispective cohort study | Methadone | Adult patients enrolled in MMT | Treatment retention | N: 1212<br>% Male: 77<br>Mean age: 42<br>Ethnicity: NA | 2 years |
| Zhou et al., 2017 (2) | China | Observational, retrospective cohort study | Methadone | Adult patients enrolled in MMT | Medication adherence | N: 10398<br>% Male: 84<br>Mean age: 38<br>Ethnicity: NA | 8 years |
| Zhu et al., 2018 | USA | Observational, retrospective study | Methadone or buprenorphine/naloxone | Opioid dependent patients | Long-term opioid abstinence (at least 5 years) | N: 699<br>% male: 65<br>Mean age: 37.4<br>Ethnicity: NA | 3 years |

Abbreviations: B-SBNT (Brief Social Behavior and Network Therapy), BCT - Behavioral couples therapy (refers to intensive therapy treatment involving once-weekly couples therapy, individual counseling and methadone for substance-abusing participants), BMT - Buprenorphine maintenance treatment, FOF - Focus on Families intervention (refers to parent skills training and case management), HIP - high intervention program of methadone plus individual counseling, CM - Contingency management, CRA - Community reinforcement approach (refers to a combination of psychoeducation, pharmacotherapy, compliance therapy, relation therapy and job counselling), LAAM - Levo-alpha-acetylmethadol harm reduction, MFT - Multiple family therapy (refers to a combination of multi-generational, structural and general supportive approaches to therapy for patients and their families), MMT - Methadone maintenance treatment, PGS - Personal Goal Setting counseling, RCT - Randomized controlled trial, SO - significant other involvement, TAU - treatment as usual.

**Table 2:** Network and Timeline Characteristics

| Author, date | Form of social network support | Network tie type | Point in treatment | Conclusions |
| --- | --- | --- | --- | --- |
| Anton et al., 1981 | Family - parents, siblings<br>Partner | Positive | During treatment | Higher retention rate for patients treated with multiple family therapy (MFT) than those who did not receive MFT. |
| Carroll et al., 2001 | Family - parent, child, or sibling (non-using)<br>Partner (non-using)<br>Peer (non-using) | Positive | During treatment | Patients who attended at least 1 family counselling session had better results for those under the significant other (SO) than those in the contingency management (CM) group with regards to retention, compliance and drug use outcomes. |
| Catalano et al., 1997 | Family - child (cohabiting) | Positive | During treatment | Parents in the experimental group (standard methadone treatment + supplemental parenting program) who had lower levels of opiate use than subject in standard methadone treatment group. |
| Catalano et al., 1999 | Family - child (cohabiting) | Positive | During and Post-treatment | Parents in experimental group who did a parent skills training and home-based case management to reduce parents' risk for relapse had lower levels of opiate use than control subjects. |
| Cerovecki et al., 2013 | Family | Negative (unstable relationship/lack of support) | Pre-treatment | Increased fatal outcome for patients with more unstable relationships and loss of continuity of care. |
| Davila Torres, 2011 | Family (cohabiting)<br>Partner (cohabiting) | Positive | Pre-treatment | Successful treatment completion associated with living in stable housing and higher perceived physician/nurse support. |
| Day et al., 2013 | Family<br>Partner<br>Peer | Positive<br>Negative (network drug use) | During treatment | Social contacts who use heroin was higher amongst patients who had used heroin in the past month, no significant effect of social contacts who supported treatment. |
| Day et al., 2018 | Family - parents, siblings, children<br>Partner<br>Peer | Positive<br>Negative (network drug use) | During and Post-treatment | No significant differences were found between the 3 intervention arms in primary or secondary outcome measures: treatment as usual (TAU, Brief Social Behavior and Network Therapy (BSBNT) ± TAU or Personal Goal Setting (PGS) ± TAU). |
| Fals-Stewart et al., 2001 | Partner (cohabiting, non-using) | Positive | Pre- and During treatment | Patients who received BCT reported greater reductions in drug use severity and family and social problems from baseline to post treatment than patients receiving standard MM treatment. |
| Fals-Stewart and O'Farrell, 2003 | Family (cohabiting, non-using)<br>Partner (cohabiting, non-using) | Negative (unstable relationship/lack of support) | Pre- During and Post-treatment | Male opioid-dependents who completed BFC with non-using family member produced better outcomes than individual therapy during treatment. |

Table 2: Network and Timeline Characteristics (continued)
| Author, date | Form of social network support | Network tie type | Point in treatment | Conclusions |
| --- | --- | --- | --- | --- |
| Feng et al., 2018 | Family (drug-using)<br>Partner (drug-using) | Positive<br>Negative (unstable relationship/lack of support AND network drug use) | Pre-treatment | Higher heroin use concurrent with treatment for patients with family members with history of heroin use. Those with family conflicts were more likely to use, while those with family who totally supported them on treatment were less likely to use. |
| Goehl et al., 1993 | Family (non-using and drug-using)<br>Partner (non-using and drug-using)<br>Peer (non-using and drug-using) | Positive<br>Negative (network drug use) | Pre-treatment | Social support was correlated with positive affect, and stress with negative affect but no measures of social support, affect, or stress correlated with the proportion of drug positive urines. However, patients with at least one drug user among the closest significant others had more positive urines than those without a drug-using significant other. |
| Gogineni et al., 2001 | Family (drug-using)<br>Partner (cohabiting, drug-using)<br>Peer (drug-using) | Positive<br>Negative (network drug use) | During treatment | Patients with substance-using live-in partners or drug-using social relationships had higher drug use, while those with emotional support had lower. |
| Grey et al., 1986 | Family<br>Partner | Positive | Pre-treatment | Drug abuse was correlated significantly with perceived family support. |
| Griffith et al., 1998 | Family<br>Partner<br>Peer (drug-using) | Positive<br>Negative (unstable relationship/lack of support AND network drug use) | Pre-treatment | A history of poor family relations was related to perceived family dysfunction and peer deviance at treatment entry; these 2 factors in turn predicted poor psychosocial functioning, which was related to higher levels of motivation. Higher motivation was associated with greater treatment engagement, which was associated with reduced opioid use and criminality. |
| Gu et al., 2013 | Family - parents, siblings, other significant others<br>Partner | Positive | During treatment | Patients who received a combination of psycho-social intervention with the standard of-care (SOC) MMT service as compared to that of the SOC MMT service alone showed lower likelihood of attrition. |
| Gu et al., 2014 | Family<br>Peer (drug-using) | Negative (unstable relationship/lack of support AND network drug use) | During treatment | Increased nonattendance at MMT clinics for patients with interpersonal conflicts with family, financial difficulty and with worry about police arrest. |
| Heinz et al., 2009 | Partner (cohabiting) | Positive | Pre-treatment | Reduced cocaine and heroin use for married participants who reported a close relationship with one's partner. |
| Hikmayani et al., 2012 | Family<br>Partner | Positive | Pre-treatment | Low retention significantly associated with absence of family support. |
| Hoang et al., 2015 | Family<br>Partner | Negative (unstable relationship/lack of support) | Pre- and During treatment | Increased continued use of heroin after MMT with likelihood of family conflict. |

Table 2: Network and Timeline Characteristics (continued)
| Author, date | Form of social network support | Network tie type | Point in treatment | Conclusions |
| --- | --- | --- | --- | --- |
| Hoang et al., 2018 | Family<br>Partner | Positive | Pre- and During treatment | Increased odds of concurrent heroin use during treatment for patients with no emotional support from family or financial stability. |
| Hojjat et al., 2017 | Partner (cohabiting) | Positive | During treatment | Harm reduction education in families of patients undergoing MMT can be effective on their marital satisfaction and treatment retention. |
| Kidorf, 2018 | Family - parents, siblings, children (non-using)<br>Partner (non-using)<br>Peer (non-using) | Positive | During treatment | Patients with drug-free community support benefited from it clinically. |
| Lee et al., 2015 | Family<br>Partner | Positive<br>Negative (unstable relationship/lack of support) | Pre-treatment | Increased expressed emotion between family members and patients is linked to retention. |
| Li et al., 2012 | Family (drug-using)<br>Partner (drug-using)<br>Peer (drug-using) | Negative (network drug use) | During treatment | Maintenance clients with substance users in their social network increase the chance of concurrent heroin use. |
| Lin et al., 2011 | Family | Positive | During treatment | Perceived family support associated with improved health and negatively correlated with concurrent substance abuse. |
| Lin et al., 2013 | Family<br>Partner | Positive | Pre- and During treatment | Perceived higher family support predicted a lower risk of MMT dropout. |
| Lister et al., 2017 | Family - parents, siblings (drug-using)<br>Partner (drug-using) | Negative (network drug use) | Pre-treatment | Close family member substance abuse associated with worse short- and long-term outcomes. |
| Lundgren et al., 2007 | Family - children (cohabiting) | Positive | Pre-treatment | Patients who resided with children, were younger and had no public health insurance were more likely to stay in MMT for 6 months or less. |
| Mutasa, 2001 | Family<br>Partner<br>Peer (drug-using) | Positive<br>Negative (unstable relationship/lack of support AND network drug use) | Pre-treatment | Medication noncompliance was associated with social company availability, family-related conflicts and peer association. |
| Nguyen et al., 2017 | Family<br>Partner | Positive | During treatment | Patients who were reminded by family members by mobile phone to adhere to treatment were more likely to do so. |
| Roozen et al., 2003 | Family<br>Partner (cohabiting)<br>Peer | Positive | During treatment | Patients who received a combination of naltrexone and CRA (community reinforced approach) had better outcomes than those with traditional treatment. |

Table 2: Network and Timeline Characteristics (continued)
| Author, date | Form of social network support | Network tie type | Point in treatment | Conclusions |
| --- | --- | --- | --- | --- |
| Rothenberg et al., 2002 | Partner | Positive | During treatment | Poorer outcomes for patients who used methadone regularly at baseline than heroin only. |
| Sarasvita et al., 2012 | Family<br>Partner<br>Peer | Positive | Pre-treatment | Patients with family support and with less peer support were less likely to drop out. |
| Shen et al., 2018 | Family - parents, children, other<br>Partner<br>Peer (non-using and drug-using) | Positive<br>Negative (network drug use) | During treatment | Patients with a spouse, child or who had a close family member were less likely to have positive urine tests for heroin. |
| Skinner et al., 2011 | Family<br>Partner<br>Peer (drug-using) | Positive<br>Negative (unstable relationship/lack of support AND network drug use) | Post-treatment | Association with drug-using friends was related to continued drug abuse. |
| Smith, 2002 | Family<br>Partner<br>Peer | Positive | Pre-treatment | Length of time in treatment was not predicted by social support, but interaction of perceived social support and orientation towards social support predict therapeutic alliance and adherence. |
| Tang, 2016 | Family<br>Partner<br>Peer | Positive | Pre-treatment | Patients were more likely to engage in sexual activity, have stronger family relationships, and experience improved health status post-treatment. |
| Torrens et al., 1996 | Family<br>Partner | Positive | Pre-treatment | Patients living with family or stable partners had higher rates of retention. |
| Tran et al., 2018 | Family<br>Partner<br>Peer | Positive<br>Negative (unstable relationship/lack of support) | During treatment | Low rate of MMT non-adherence associated with job stability, engagement in self-care, and active participation of partners, family and friends in the treatment process. |
| Wasserman et al., 2001 | Family<br>Partner<br>Peer (non-using and drug-using) | Positive<br>Negative (network drug use) | During treatment | Having social networks with less drug users (abstinence specific structural support) correlated with reduced drug use and demoralization for cocaine abstinence but not opiate abstinence. There was no effect for general support. |
| Yandoli et al., 2002 | Family<br>Partner | Positive | During treatment | Family therapy produced significantly more drug-free subjects than standard treatment at six months and at twelve months. |
| Yang et al., 2013 | Family<br>Partner | Positive<br>Negative (unstable relationship/lack of support) | Pre-treatment | Protective factors for MMT retention were: strong relationships with family, living on support of family or friends and not communicating with former drug taking peers in the past month. |

Table 2: Network and Timeline Characteristics (continued)
| Author, date | Form of social network support | Network tie type | Point in treatment | Conclusions |
| --- | --- | --- | --- | --- |
| Zhou et al., 2017<br>(1) | Family<br>Partnee<br>Peer | Positive | Pre-treatment | Patients with poor social support are more likely to terminate treatment. |
| Zhou et al., 2017<br>(2) | Family (cohabiting)<br>Partner (cohabiting)<br>Peer (drug-using) | Positive<br>Negative (network drug use) | Pre-treatment | Patients who had no contact with peer drug users had a lower dropout risk. |
| Zhu et al., 2018 | Family<br>Partner<br>Peer | Positive | Post-treatment | Long-term abstinence was positively associated with greater social support. |

Abbreviations: BMT - Buprenorphine maintenance treatment, CM - Contingency management, LAAM - Levo-alpha-acetylmethadol harm reduction, MMT - Methadone maintenance treatment, RCT - Randomized controlled trial

**Figure 2:**
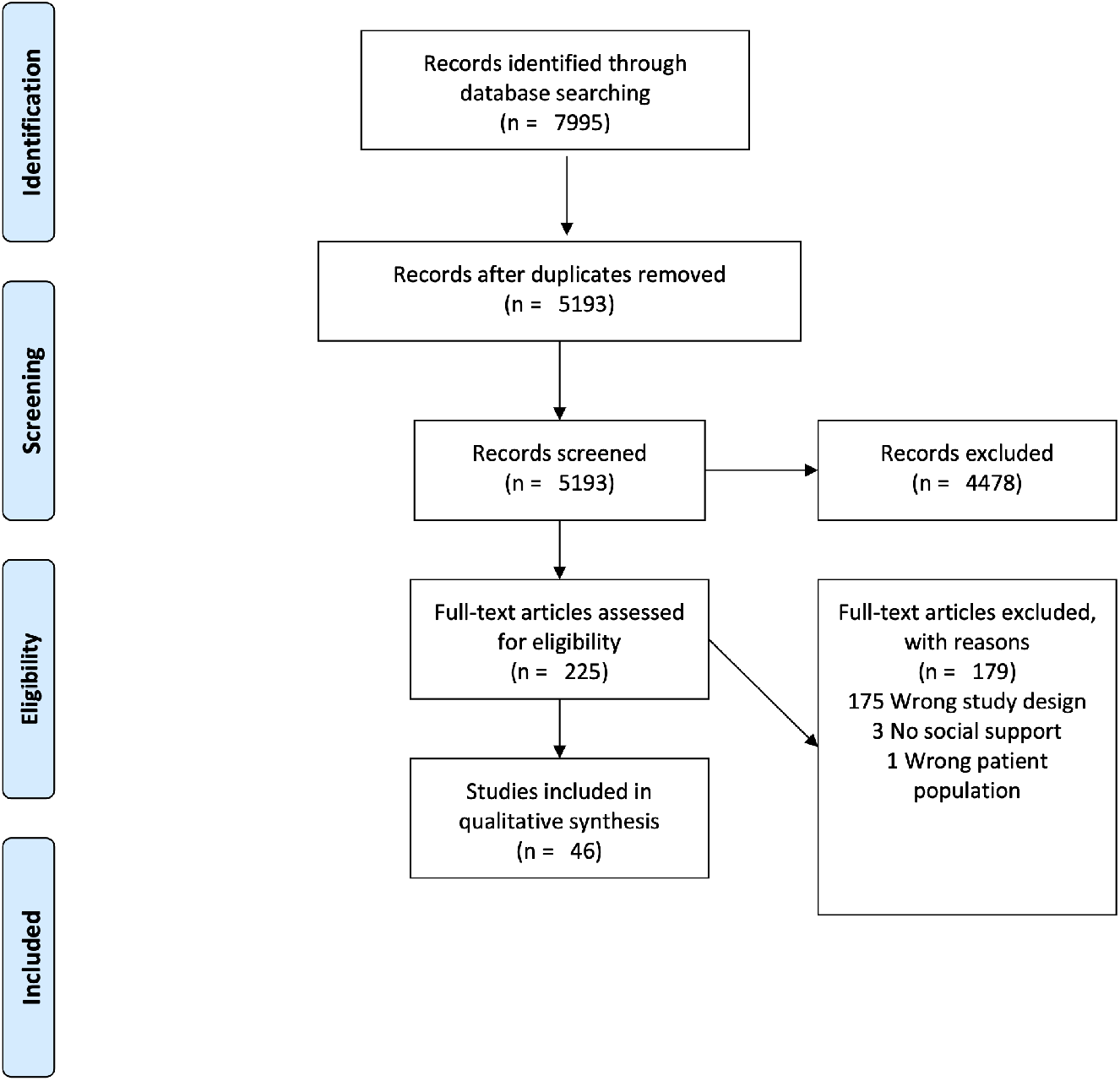
PRISMA flow diagram. Preferred Reporting Items for Systematic Review and Meta-Analysis (PRISMA) flow diagram of study selection, detailing stages of selection, exclusion, and review.

### Quality assessments

Tables 3 and 4 indicated the quality of experimental and observational studies. For experimental studies, allocation concealment was rarely reported and its impact on bias was not clear. Impact of bias referred to the impact of the quality categories in Table 3 on the administration of the experimental method and therefore was not completed for observational studies. The quality ratings for observational studies were high overall. Relevant evidence and statistical significance for observational (see Fig 3) and experimental studies (see Fig 4) were indicated with a harvest plot [45]. Thirty observational studies met all five criteria. Three observational studies met four criteria. Of observational studies assessing social network support pre-treatment, nineteen met five criteria and two met four criteria. Eleven studies assessing social network support during treatment and three studies assessing social support post-treatment met five criteria, respectively. Two studies assessing social support pre-treatment and two studies during treatment met four criteria. No experimental studies met all five criteria. Two experimental studies that assessed social network support during treatment and a single post-treatment met four criteria. The remaining studies in this review met between zero and three criteria.

**Table 3:**
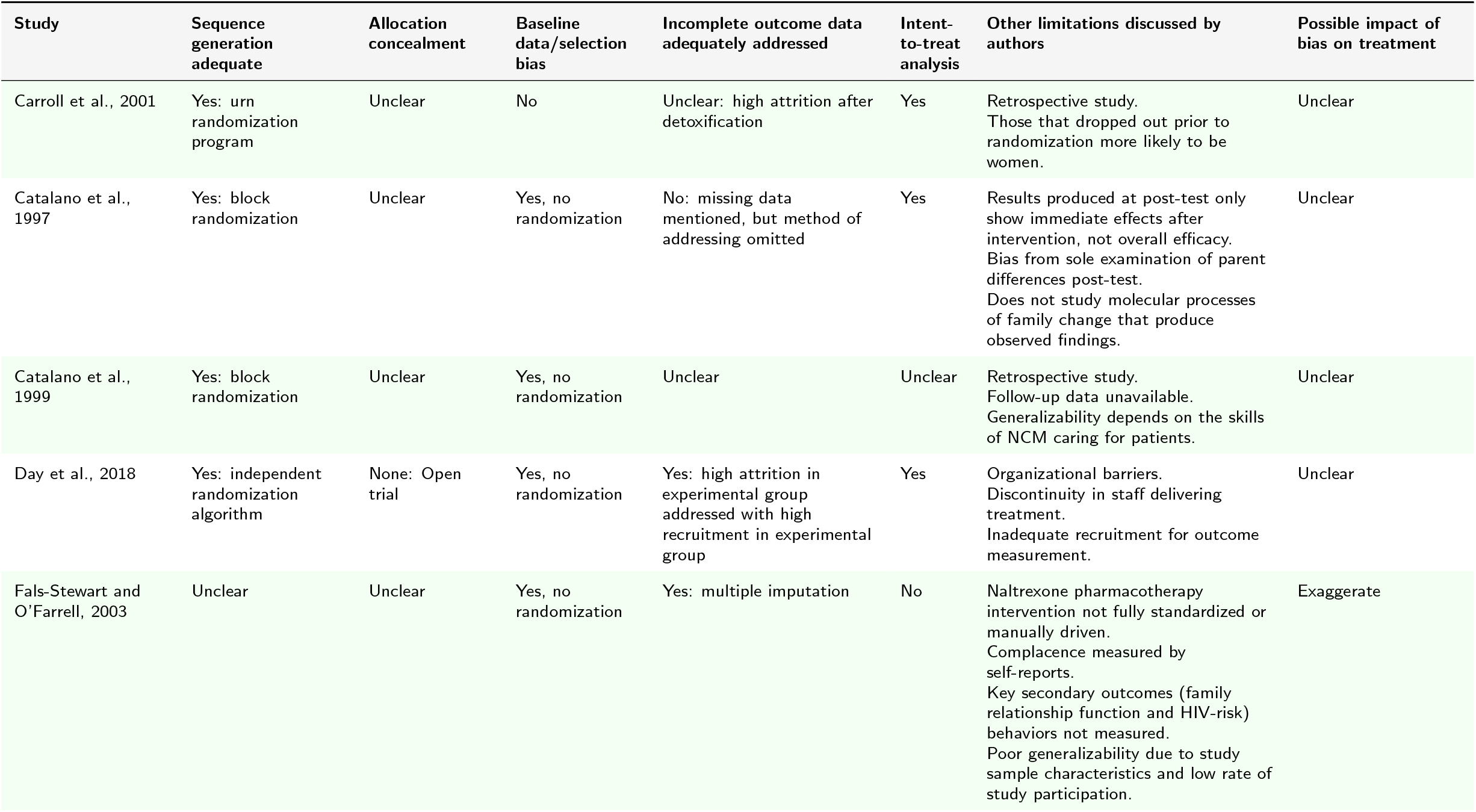
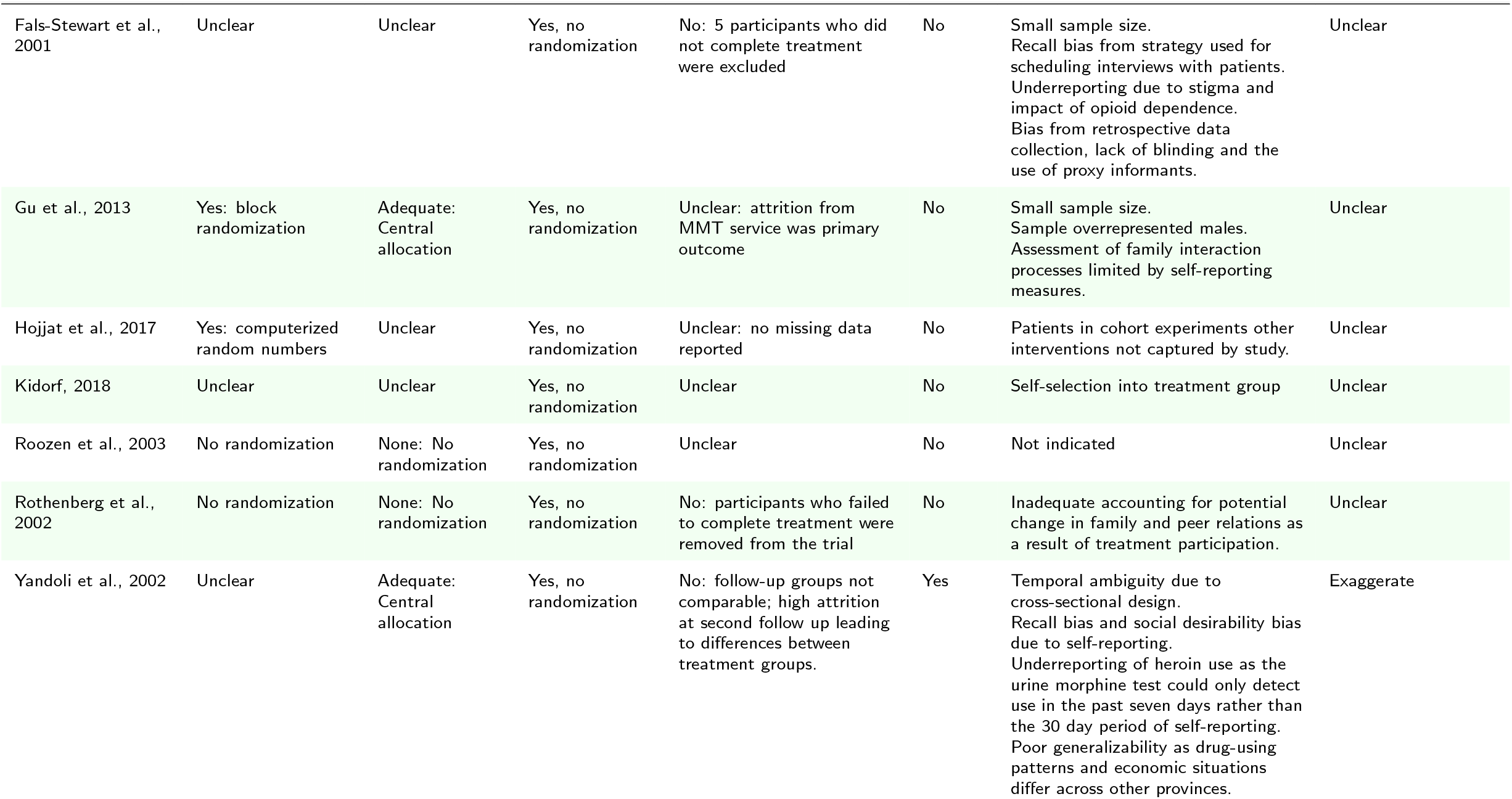
Quality of Experimental Studies

| Study | Sequence generation adequate | Allocation concealment | Baseline data/selection bias | Incomplete outcome data adequately addressed | Intent-to-treat analysis | Other limitations discussed by authors | Possible impact of bias on treatment |
| --- | --- | --- | --- | --- | --- | --- | --- |
| Carroll et al., 2001 | Yes: urn randomization program | Unclear | No | Unclear: high attrition after detoxification | Yes | Retrospective study. Those that dropped out prior to randomization more likely to be women. | Unclear |
| Catalano et al., 1997 | Yes: block randomization | Unclear | Yes, no randomization | No: missing data mentioned, but method of addressing omitted | Yes | Results produced at post-test only show immediate effects after intervention, not overall efficacy. Bias from sole examination of parent differences post-test. Does not study molecular processes of family change that produce observed findings. | Unclear |
| Catalano et al., 1999 | Yes: block randomization | Unclear | Yes, no randomization | Unclear | Unclear | Retrospective study. Follow-up data unavailable. Generalizability depends on the skills of NCM caring for patients. | Unclear |
| Day et al., 2018 | Yes: independent randomization algorithm | None: Open trial | Yes, no randomization | Yes: high attrition in experimental group addressed with high recruitment in experimental group | Yes | Organizational barriers. Discontinuity in staff delivering treatment. Inadequate recruitment for outcome measurement. | Unclear |
| Fals-Stewart and O'Farrell, 2003 | Unclear | Unclear | Yes, no randomization | Yes: multiple imputation | No | Naltrexone pharmacotherapy intervention not fully standardized or manually driven. Complacency measured by self-reports. Key secondary outcomes (family relationship function and HIV-risk) behaviors not measured. Poor generalizability due to study sample characteristics and low rate of study participation. | Exaggerate |

Table 3: Quality of Experimental Studies (continued)
| Study | Sequence generation adequate | Allocation concealment | Baseline data/selection bias | Incomplete outcome data adequately addressed | Intent-to-treat analysis | Other limitations discussed by authors | Possible impact of bias on treatment |
| --- | --- | --- | --- | --- | --- | --- | --- |
| Fals-Stewart et al., 2001 | Unclear | Unclear | Yes, no randomization | No: 5 participants who did not complete treatment were excluded | No | Small sample size. Recall bias from strategy used for scheduling interviews with patients. Underreporting due to stigma and impact of opioid dependence. Bias from retrospective data collection, lack of blinding and the use of proxy informants. | Unclear |
| Gu et al., 2013 | Yes: block randomization | Adequate: Central allocation | Yes, no randomization | Unclear: attrition from MMT service was primary outcome | No | Small sample size. Sample overrepresented males. Assessment of family interaction processes limited by self-reporting measures. | Unclear |
| Hojjat et al., 2017 | Yes: computerized random numbers | Unclear | Yes, no randomization | Unclear: no missing data reported | No | Patients in cohort experiments other interventions not captured by study. | Unclear |
| Kidorf, 2018 | Unclear | Unclear | Yes, no randomization | Unclear | No | Self-selection into treatment group | Unclear |
| Roizen et al., 2003 | No randomization | None: No randomization | Yes, no randomization | Unclear | No | Not indicated | Unclear |
| Rothenberg et al., 2002 | No randomization | None: No randomization | Yes, no randomization | No: participants who failed to complete treatment were removed from the trial | No | Inadequate accounting for potential change in family and peer relations as a result of treatment participation. | Unclear |
| Yandoli et al., 2002 | Unclear | Adequate: Central allocation | Yes, no randomization | No: follow-up groups not comparable; high attrition at second follow up leading to differences between treatment groups. | Yes | Temporal ambiguity due to cross-sectional design. Recall bias and social desirability bias due to self-reporting. Underreporting of heroin use as the urine morphine test could only detect use in the past seven days rather than the 30 day period of self-reporting. Poor generalizability as drug-using patterns and economic situations differ across other provinces. | Exaggerate |

**Table 4:**
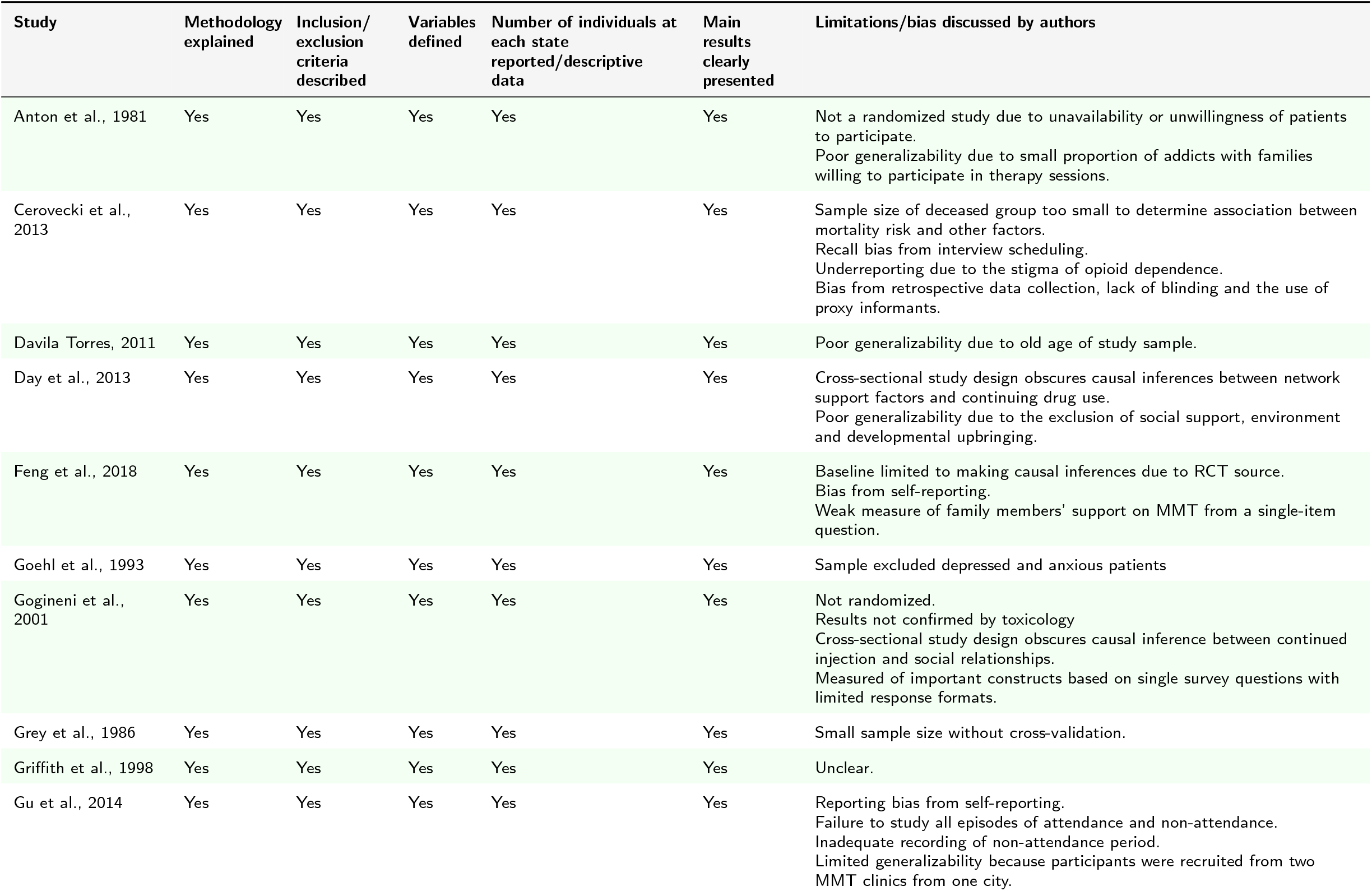
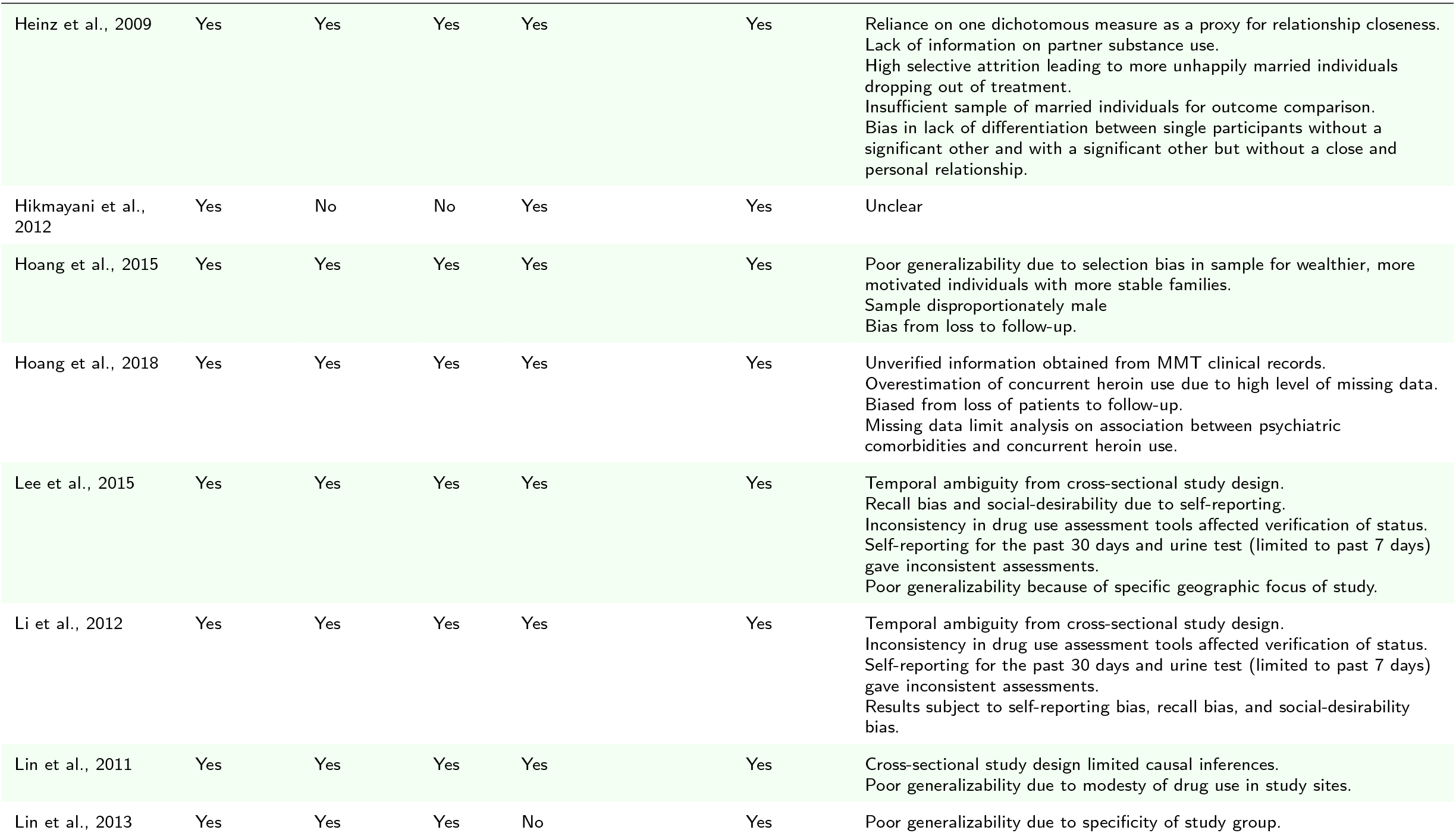
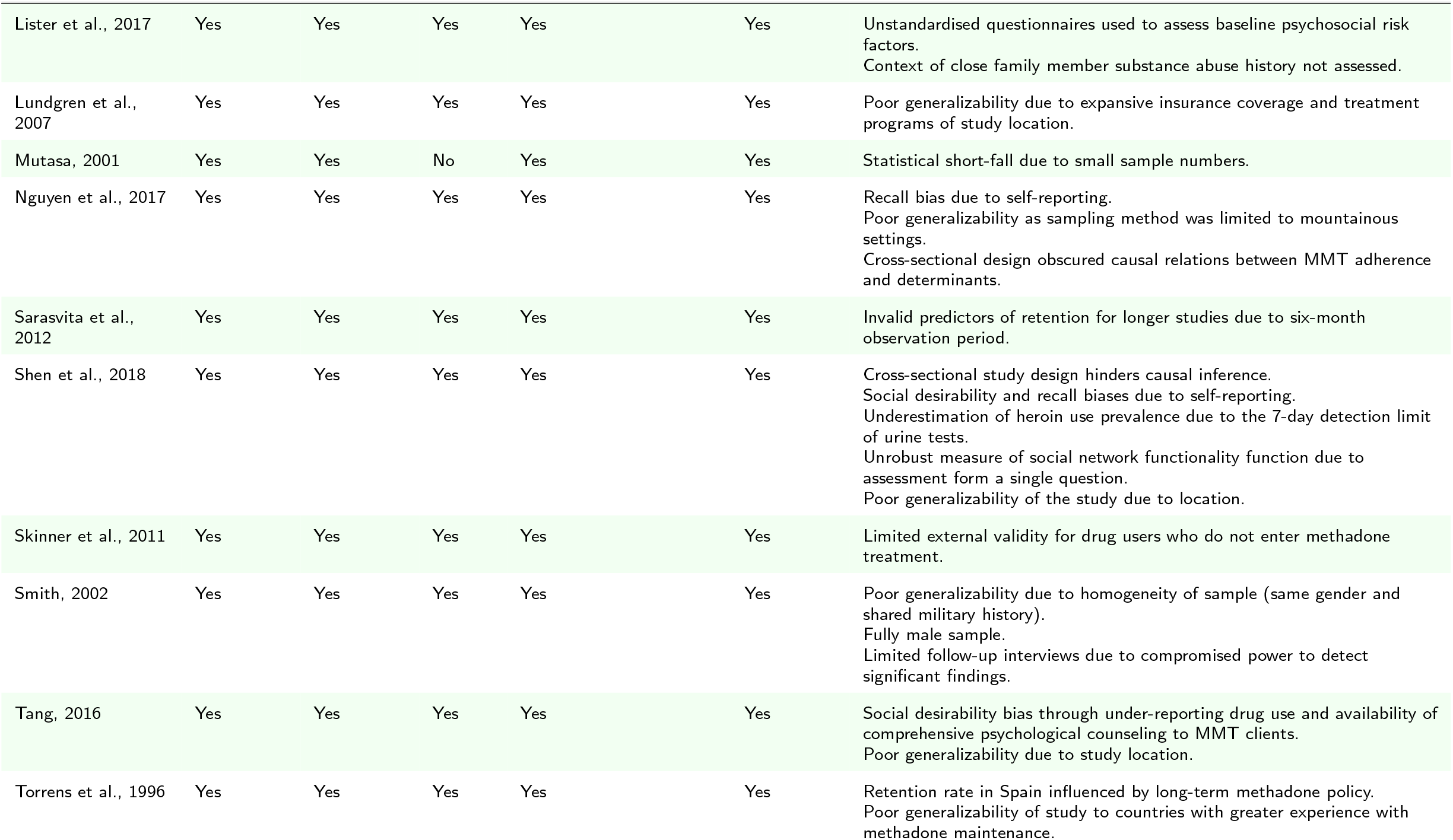
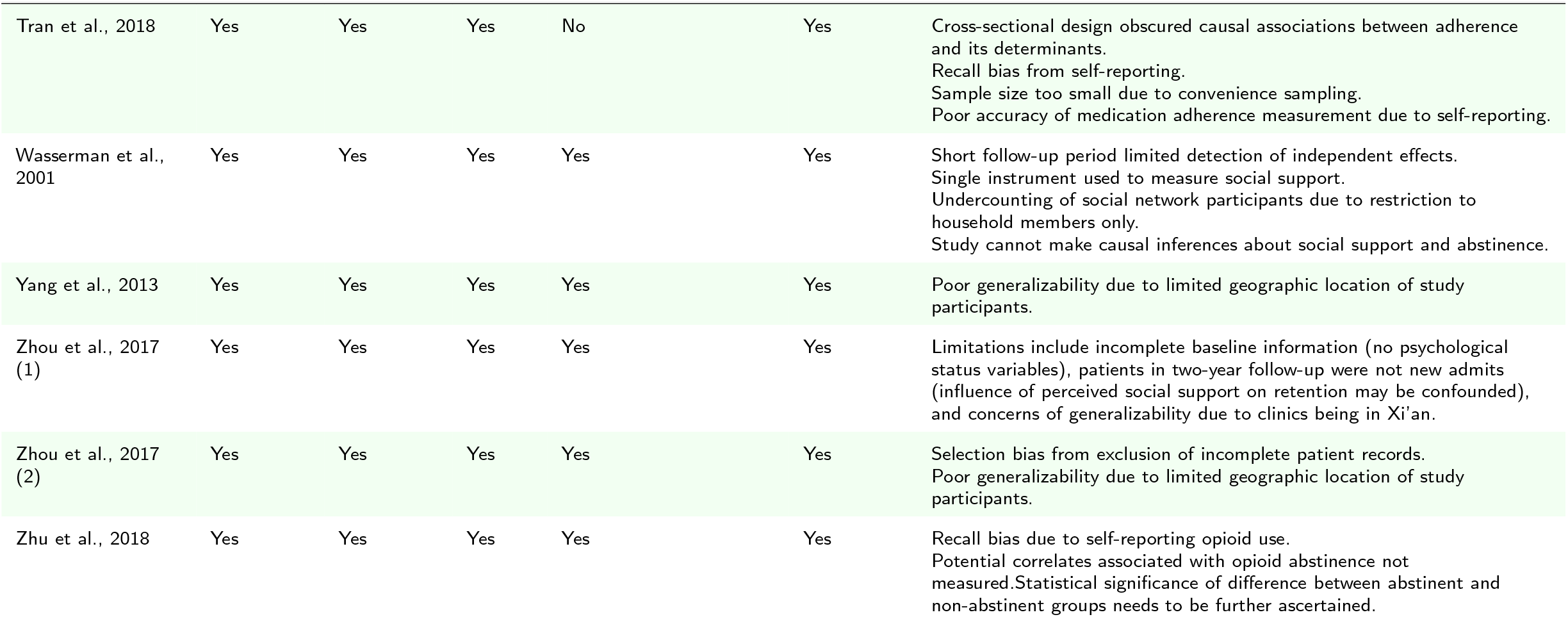
Quality of Observational Studies

**Figure 3:**
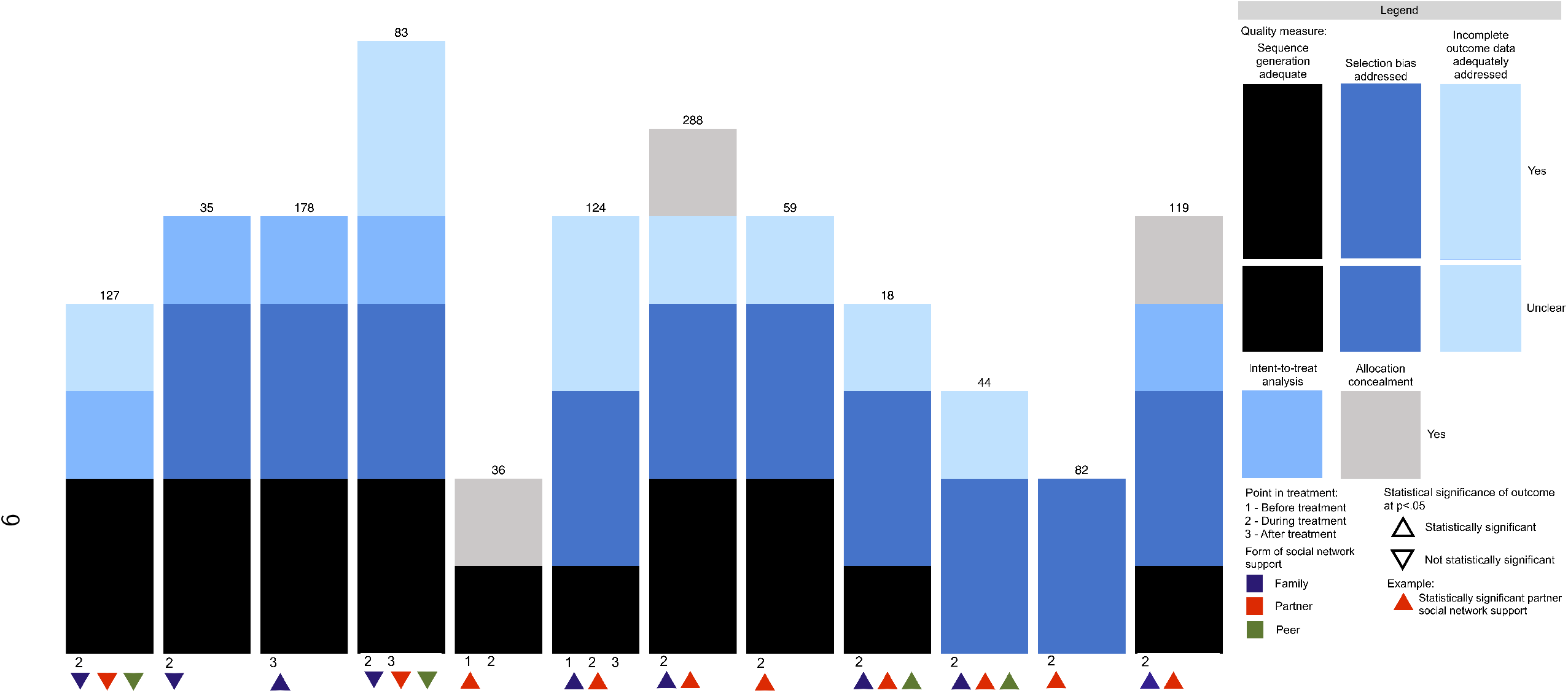
Observational study harvest plot. Evidence for observational studies regarding the role of social network support on treatment outcomes for medication for opioid use disorder. Each study falls into a category represented by a stacked bar. The height of each component corresponds to a quality score representing the suitability of study design with respect to five quality measures: description of inclusion/exclusion criteria, clear presentation of main results, explanation of methodology, presence of descriptive data and definition of variables. Each bar is annotated with the sample size, statistical significance of outcome, and form of social network support.

**Figure 4:**
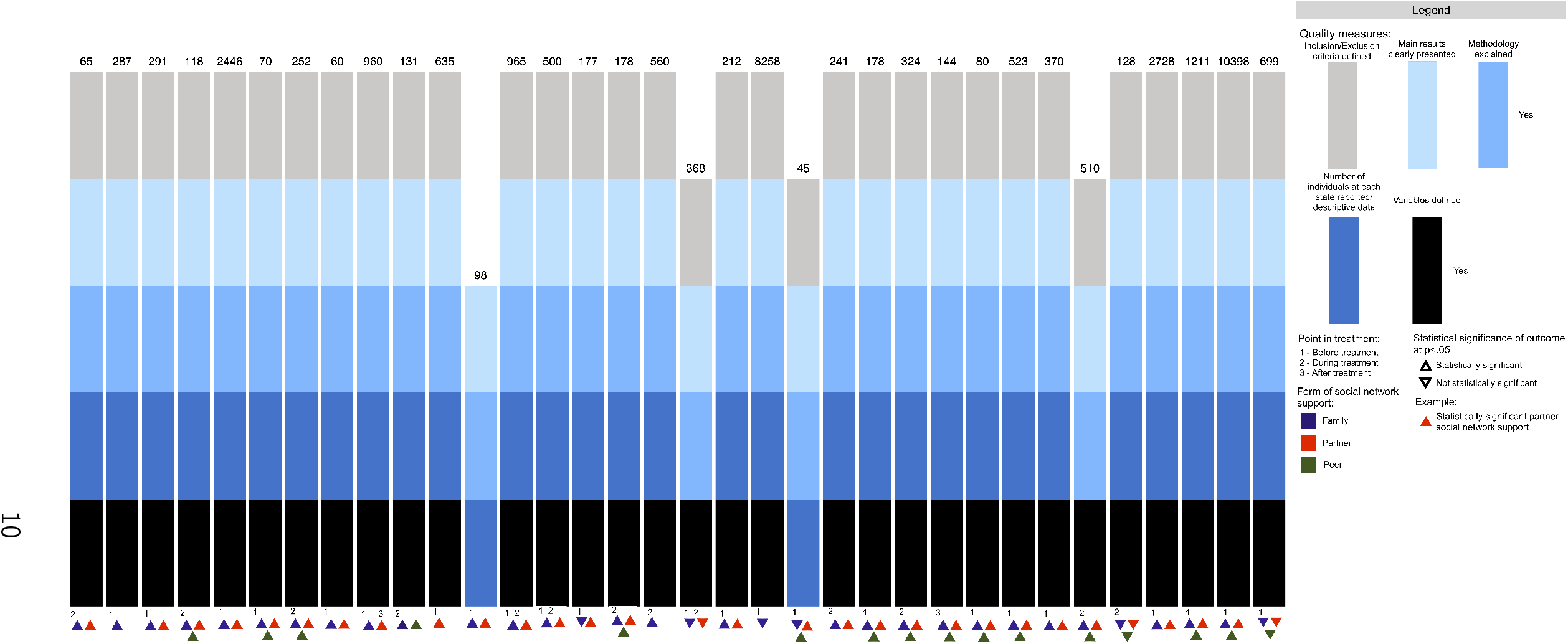
Experimental study harvest plot. Evidence for experimental studies regarding the role of social network support on treatment outcomes for medication for opioid use disorder.Each study falls into a category represented by a stacked bar. The height of each component corresponds to a quality score representing the suitability of study design with respect to five quality measures: allocation concealment, addressing of incomplete outcome data, intent-to-treat analysis, addressing of selection bias and adequate sequence generation. Each bar is annotated with the sample size, statistical significance of outcome, and form of social network support. Studies without above quality measures are represented with no bar.

### Pre-treatment

Studies characterized as assessing social network characteristics pre-treatment were often studies which gathered baseline survey information about their participants, typically at treatment initiation. Information at this point in the treatment timeline is of particular importance for two reasons: first, because it reveals patterns in the social network composition of individuals entering MOUD programs, and second, because levels of support at treatment initiation may be predictive of treatment success at later time points. Twenty-four studies assessed participant social network support pre-treatment. Two were experimental and 22 were observational. Nineteen studies indicated that pre-treatment-assessed social network ties were significantly related to MOUD treatment outcomes, while two studies had non-significant results and three studies had mixed results.

#### Family social network support

Seventeen of 22 studies exploring family social network ties pre-treatment demonstrated statistically significant relationships with treatment outcomes. Among the observational studies, many evaluated self-reports of perceived family support [46, 47, 48, 49] or cohabitation with family members [50, 51]. For example, one study (N=2728) found that patients who lived with family or who had good relationships with their family were more likely to be retained in treatment for a longer period than those that did not [52]. Other studies noted the negative effects of having family conflicts [53], perceived criticism from family [54], or contact with drug-using family members [55] at baseline. One study (N=2446) analyzing Addiction Severity Index items noted that those who reported having family problems or having family members who used heroin were more likely to use heroin concurrently during treatment [46]. Another study (N=435) that utilized a specific effects model to link pre-treatment social factors, engagement during treatment, and post-treatment outcomes found significant correlations between perceived family dysfunction and peer drug-use at intake [44]. Interestingly, the authors’ model supported the idea that family dysfunction and peer deviance were related to lower psychosocial functioning which was related to greater engagement in treatment––greater treatment engagement predicted a lower number of days of drug-use one-year after treatment termination.

#### Partner social network support

Twenty-two studies explored partner social network support and 18 demonstrated statistically significant relationships with treatment outcomes. In general, studies that explored partner relationships were often classified as assessing family relationships as well, so the classification results have significant overlap. One observational study (N=635) which looked at partner-specific support found that marriage predicted fewer heroin-positive urine samples during treatment and that high perceived closeness to partners among married participants predicted even fewer positive urine screens [56].

#### Peer social network support

Eight studies explored peer social network support, and all were observational studies. Six demonstrated statistically significant relationships with treatment outcomes, while two studies had mixed results. Common themes included analysis of respondent’s perceived support from friends, cohabitating with non-family friends, and having less contact with drug-using peers. One study (N=1212) which assessed the peer component of the Multidimensional Scale of Perceived Social Support found that greater perceived peer support at intake, even when controlling for family support and family relationship quality, predicted longer program retention [57]. Another study (N=10398) noted that reports of having no contact with peer drug users in last month significantly predicted longer retention [58]. Of the studies with significant results, only one (N=178) reported that perceived peer support as assessed by the Community Assessment Inventory was associated with a higher likelihood of dropout [59].

### During treatment

Studies characterized as assessing social network support during treatment were either those that gathered self-report information about participant social ties after treatment had already begun or those that actively involved participant social ties in treatment. All experimental studies inherently had at least one during-treatment component to be included in the review. Twenty-five studies assessed participant social network support during treatment. Twelve were experimental and 13 were observational. Nineteen studies indicated that during-treatment-assessed social network support was significantly associated with MOUD treatment outcomes, while six studies were non-significant. Nine of the 12 RCTs that involved social network support in MOUD treatment showed improved outcomes.

#### Family social network support

Seventeen out of 22 studies exploring family social network support during treatment demonstrated statistically significant relationships with treatment outcomes. Improvements in treatment outcome were found in both experimental and observational studies. One experimental study (N=119) explored the efficacy of family therapy versus standard treatment involving methadone maintenance and individual counseling, finding that there was a significantly larger number of drug-free participants in the treatment group at both 6- and 12-month follow-ups [60]. Family support in this case included attendance to biweekly therapy sessions which emphasized rebuilding relationships and methadone reduction. Still, another experimental study randomly employed Brief Social Behavior and Network Therapy––which allows MOUD patients to map their social network of significant others and invite them to treatment sessions––found no significant difference in number of days abstinent from heroin between the treatment group (N=26), control (N=30), and individual therapy groups (N=27) at three or 12 month follow-ups [41]. It should be noted that this study recorded particularly low attendance to treatment sessions (44% attendance), which continues to be a primary obstacle for psychosocial interventions in many clinical settings.

Observational studies in the during treatment group reported similar findings to those in the pre-treatment group, citing improvements in treatment outcomes related to perceived social support during treatment [40, 61, 62] and having family members remind the patient to take medication [63], while risk factors for unsuccessful treatment were experiencing family conflicts and maintaining contact with drug-using family members during treatment [40, 64, 65].

#### Partner social network support

Seventeen of 21 studies exploring partner social network support during treatment noted a significant relationship with treatment outcomes. In general, studies which explored partner relationships were often classified as assessing family relationships as well, so the classification results have significant overlap. However, three experimental studies looked specifically at the impact of partner-specific involvement in treatment. One experimental study randomly employed Behavioral Couples Therapy centered around partners practicing positive reinforcement of abstinence, finding that those in the couples therapy condition (N=19) had fewer positive urine screens during treatment than those in the individual therapy condition (N=17) [66]. Another study (N=47) cited improvements in retention and percentage opiate-free urine screens among a group receiving couples therapy [67], while the last study (N=48) which utilized a randomized intervention to educate spouses about the harm reduction approach found a significant improvement in marital satisfaction but no significant difference in retention at one month or six months [68].

#### Peer social network support

Seven of the 10 studies which explored peer social support during treatment found a significant association with treatment outcomes. All 10 studies which included assessments of peer support also included family or partner support, so the classification results have significant overlap. Some studies noted the general positive impact of perceived support from friends or non-using friend involvement in treatment [43, 69], while one observational study (N=510) in particular noted that patients who disclosed their health status to friends rather than family missed a significantly lower number of MOUD doses [70]. Most studies that assessed peer relationship information did so in the context of peer drug use and found a negative impact of maintaining contact with peers in the drug-using network during treatment on outcomes [61, 64, 71, 41, 42].

### Post-treatment

Studies characterized as assessing social network support post-treatment were those that elicited information from study participants with a follow-up that lasted beyond the end of treatment termination. Studies that assessed post-treatment social support were of particular importance, as there are gaps in the understanding of patient social environments after treatment termination despite high rates of attrition and high rates of repeated treatment attempts in MOUD programs. Five studies assessed participant social network support post-treatment. Three were experimental and two were observational. Three studies indicated that post-treatment-assessed social network support was significantly associated with MOUD treatment outcomes, while one study found non-significant associations and one study reported mixed effects.

#### Family social network support

All of the post-treatment studies explored at least one form of family social ties. Improvements in treatment outcomes were found in both experimental and observational studies. In one experimental study, parents in MOUD were randomly assigned to the Focus on Families intervention––a curriculum that combines parent skills training and home-based case management––and given a follow-up interview within a month of the program termination. The authors noted that parents in the intervention group (N=82) had significantly more family discussions regarding their drug use, better relapse coping skills, and a lower frequency of opiate use at follow-up compared to the control group (N=62) [72]. Another observational study completed a 12-year follow-up with the same cohort of 144 participants in the Focus on Families intervention, finding generally high rates of incarceration, residential displacement, and health problems among all participants. Importantly, the authors noted that the small proportion of participants who maintained recovery (i.e., continued abstinence, no incarceration, clean urinalysis) for at least 5 years were predicted by having less relationship disruptions (e.g., less changes in marital status, maintaining relationships with children)[73].

#### Partner social network support

Four of the five post-treatment studies also included assessment of partner social support, so the classification results have significant overlap. One of the previously mentioned experimental studies which included a randomly assigned Behavioral Family Counseling component of which a high proportion of participants completed with their partner also elicited post-treatment information of participants at a one-year follow-up. The authors noted that both the treatment and control groups in the study saw significant decreases in percentage of days abstinent and length of continuous abstinence that year, that both groups also had significant improvements in the social and family functioning component of the Addiction Severity Index at follow-up, and that those in the treatment group (N=62) had significantly better outcomes and social/family functioning at follow-up than the control group (N=62) [74].

#### Peer social network support

Three out of five post-treatment studies assessed a component of peer social network support, so the classification results have significant overlap. An aforementioned study (N=144) which investigating a 12-year follow-up of the Focus on Families cohort found that, in addition to less family relationship disruptions, recovery for at least five years was also negatively associated with having someone who actively used illicit substances among one’s four closest friends [73]. Another observational study (N=1269) which interviewed participants about their perceived social support from family and friends at least five years after MOUD termination found that those those with higher perceived social support at the time of interview were also more likely to be heroin abstinent [75].

### Negative and positive social connections

Twenty-seven studies investigated positive social network support, seven studies investigated negative social network ties, and 13 studies investigated both positive and negative social ties. Of the studies that assessed an aspect of positive social support, 36 included family ties, 35 included partner ties, and 22 included peer ties. Of the studies that assessed an aspect of negative social ties, 20 included family ties, 19 included partner ties, and 14 included peer ties.

Of the studies that included negative social ties, there was a large cluster that included specific discussion of the negative influence of maintaining peer ties in the drug-using network. For example, one observational study (N=118) which utilized the Important People Drug and Alcohol interview to map egocentric patient networks found a weak positive impact of having network members that actively supported treatment and a strong negative impact of having network members that used heroin with respect to respondent’s heroin abstinence in the previous month [40].

### Synthesis

Table 5 synthesized included studies and indicated if there were statistically significant findings or no effect. Table 5 also indicated whether biases may have understated or over-reported treatment effects, if any. Evidence was not consistent for all points at which social network support was assessed, although studies overall indicated that social network support, or lack thereof, was statistically significantly associated with MOUD treatment outcomes. This information was derived from Tables 3 and 4. For experimental studies, bias was considered likely to understate positive outcomes in 0 studies, to exaggerate in two and unclear in the remaining studies. The most common source of bias for experimental studies was lack of intent-to-treat analysis.

**Table 5:** Synthesis

| Study | Study type | Statistical significance of outcome at $p < .05$ | Form of social network support | Point in treatment timeline that social network is assessed | MOUD drug used | Intervention (if any) |
| --- | --- | --- | --- | --- | --- | --- |
| Anton et al., 1981 | Observational | Statistically significant | Family - parents, siblings<br>Partner | During treatment | Naltrexone | Multiple family therapy (no randomization, self-elected)<br>Standard: naltrexone + individual counseling |
| Carroll et al., 2000 | Experimental | No effect | Family - parent, child, or sibling (non-using)<br>Partner (non-using)<br>Peer (non-using) | During treatment | Naltrexone | Significant other involvement +<br>Contingency management +<br>standard treatment<br>Contingency management +<br>standard treatment<br>Standard: Naltrexone + treatment<br>Cognitive Behavioral Therapy in group setting |
| Catalano et al., 1997 | Experimental | Statistically significant | Family - child (cohabiting) | During and Post-treatment | Methadone | Family counseling (Focus on Families intervention: parent skills training and home-based case management service)<br>Standard: not specified |
| Catalano et al., 1999 | Experimental | Statistically significant | Family - child (cohabiting) | During treatment | Methadone | Family counseling (Focus on Families intervention: parent skills training and home-based case management service)<br>Standard: not specified |
| Cerovecki et al., 2013 | Observational | Statistically significant | Family | Pre-treatment | Methadone | Standard: not specified |
| Davila Torres, 2011 | Observational | Statistically significant | Family (cohabiting)<br>Partner (cohabiting) | Pre-treatment | Methadone | Standard: MMT |
| Day et al., 2013 | Observational | Statistically significant [Positive (family, partner, peer)]<br>Statistically significant [Negative (family, partner, peer)] | Family<br>Partner<br>Peer | During treatment | Methadone or buprenorphine | Standard: not specified |

Table 5: Synthesis (continued)
| Study | Study type | Statistical significance of outcome at $p < .05$ | Form of social network support | Point in treatment timeline that social network is assessed | MOUD drug used | Intervention (if any) |
| --- | --- | --- | --- | --- | --- | --- |
| Day et al., 2018 | Experimental | No effect [Positive (family, partner, peer)]<br>No effect [Negative (family, partner, peer)] | Family - parents, siblings, children<br>Partner<br>Peer | During and Post-treatment | Methadone or buprenorphine | Standard + Brief Social Behavior and Network Therapy (B-SBNT)<br>Standard + Personal Goal Setting (PSG)<br>Standard: individual case management |
| Fals-Stewart et al., 2001 | Experimental | Statistically significant | Partner (cohabiting, non-using) | Pre- and During treatment | Methadone | Behavioral Couples Therapy + standard treatment<br>Standard: MMT + individual counseling |
| Fals-Stewart and O'Farrell, 2003 | Experimental | Statistically significant | Family (cohabiting, non-using)<br>Partner (cohabiting, non-using) | Pre-, During and Post-treatment | Naltrexone | Behavioral family counseling + standard treatment<br>Standard: individual based therapy (individual counseling, group counseling) |
| Feng et al., 2018 | Observational | Statistically significant [Positive (family, partner)]<br>Statistically significant [Negative (family, partner)] | Family (drug-using)<br>Partner (drug-using) | Pre-treatment | Methadone | Standard: MMT |
| Goehl et al., 1993 | Observational | No effect [Positive (family, partner, peer)]<br>Statistically significant [Negative (family, partner, peer)] | Family (non-using and drug-using)<br>Partner (non-using and drug-using)<br>Peer (non-using and drug-using) | Pre-treatment | Methadone | Standard : MMT |
| Gogineni et al., 2000 | Observational | Statistically significant | Family (drug-using)<br>Partner (cohabiting, drug-using)<br>Peer (drug-using) | During treatment | Methadone | Standard: not specified |
| Grey et al., 1986 | Observational | Statistically significant | Family<br>Partner | Pre-treatment | Methadone or naltrexone | Standard: not specified |

Table 5: Synthesis (continued)
| Study | Study type | Statistical significance of outcome at $p < .05$ | Form of social network support | Point in treatment timeline that social network is assessed | MOUD drug used | Intervention (if any) |
| --- | --- | --- | --- | --- | --- | --- |
| Griffith et al., 1998 | Observational | Statistically significant [Positive (family, partner)]<br>Statistically significant [Negative (family, peer)] | Family<br>Partner<br>Peer (drug-using) | Pre- treatment | Methadone | Standard: MMT with individual and group counseling |
| Gu et al., 2013 | Experimental | Statistically significant | Family - parents, siblings<br>Partner | During treatment | Methadone | Behavioral maintenance therapy + standard treatment<br>Standard: MMT (no counseling) |
| Gu et al., 2014 | Observational | Statistically significant | Family<br>Peer (drug-using) | During treatment | Methadone | Standard: not specified |
| Heinz et al., 2009 | Observational | Statistically significant | Partner (cohabiting) | Pre-treatment | Methadone | Standard: MMT + contingency management + individual counseling |
| Hikmayani et al., 2012 | Observational | Statistically significant | Family<br>Partner | Pre-treatment | Methadone | Standard: MMT |
| Hoang et al., 2015 | Observational | Statistically significant | Family<br>Partner | Pre- and During treatment | Methadone | Standard: not specified |
| Hoang et al., 2018 | Observational | Statistically significant | Family<br>Partner | Pre- and During treatment | Methadone | Standard: not specified |
| Hojjat et al., 2016 | Experimental | No effect | Partner (cohabiting) | During treatment | Methadone | Standard + Group training<br>Standard: MMT |
| Kidorf, 2018 | Experimental | Statistically significant | Family - parents, siblings, children (non-using)<br>Partner (non-using)<br>Peer (non-using) | During treatment | Methadone | Social network activation group therapy + standard treatment<br>Standard: MMT + individual and group counselling + community activity (mutual peer-support groups or church) |
| Lee et al., 2015 | Observational | No effect [Positive (family, partner)]<br>Statistically significant [Negative (family, partner)] | Family<br>Partner | Pre-treatment | Methadone | Standard: MMT |

Table 5: Synthesis (continued)
| Study | Study type | Statistical significance of outcome at $p < .05$ | Form of social network support | Point in treatment timeline that social network is assessed | MOUD drug used | Intervention (if any) |
| --- | --- | --- | --- | --- | --- | --- |
| Li et al., 2012 | Observational | Statistically significant | Family (drug-using)<br>Partner (drug-using)<br>Peer (drug-using) | During treatment | Methadone | Standard: MMT |
| Lin et al., 2011 | Observational | No effect | Family | During treatment | Methadone | Standard: MMT |
| Lin et al., 2013 | Observational | No effect | Family<br>Partner | Pre- and During treatment | Methadone | Standard: MMT |
| Lister et al., 2017 | Observational | Statistically significant | Family - parents, siblings (drug-using)<br>Partner (drug-using) | Pre-treatment | Methadone | Standard: MMT |
| Lundgren et al., 2007 | Observational | No effect | Family - children (cohabiting) | Pre-treatment | Methadone | Standard: MMT |
| Mutasa, 2001 | Observational | No effect [Positive (family, partner, peer)]<br>Statistically significant [Negative (family, partner, peer)] | Family<br>Partner<br>Peer (drug-using) | Pre-treatment | Methadone | Standard: MMT |
| Nguyen et al., 2017 | Observational | Statistically significant | Family<br>Partner | During treatment | Methadone | Standard: not specified |
| Roizen et al., 2003 | Experimental | Statistically significant | Family<br>Partner (cohabiting)<br>Peer | During treatment | Methadone or naltrexone | Naltrexone + CRA (with social network emphasis)<br>Standard: MMT |
| Rothenberg et al., 2002 | Experimental | Statistically significant | Partner | During treatment | Naltrexone | Standard: Behavioral Naltrexone Therapy (naltrexone + network therapy + CRA) |
| Sarasvita et al., 2012 | Observational | Statistically significant | Family<br>Partner<br>Peer | Pre-treatment | Methadone | Standard: MMT |
| Shen et al., 2018 | Observational | Statistically significant [Positive (family, partner, peer)]<br>Statistically significant [Negative (family, partner, peer)] | Family - parents, children, other<br>Partner<br>Peer (non-using and drug-using) | During treatment | Methadone | Standard: MMT |

Table 5: Synthesis (continued)
| Study | Study type | Statistical significance of outcome at $p < .05$ | Form of social network support | Point in treatment timeline that social network is assessed | MOUD drug used | Intervention (if any) |
| --- | --- | --- | --- | --- | --- | --- |
| Skinner et al., 2011 | Observational | No effect [Positive (family, partner)]<br>Statistically significant [Negative (partner, peer)] | Family<br>Partner<br>Peer (drug-using) | Post-treatment | Methadone | Standard: not specified (10 year follow up to original Focus on Families study, which included parent counseling and home-based case management for parents with OUD) |
| Smith, 2002 | Observational | Statistically significant | Family<br>Partner<br>Peer | Pre-treatment | Methadone | Standard: MMT |
| Tang, 2016 | Observational | Statistically significant | Family<br>Partner<br>Peer | Pre-treatment | Methadone | Standard: MMT |
| Torrens et al., 1996 | Observational | Statistically significant | Family<br>Partner | Pre-treatment | Methadone | Standard: MMT |
| Tran et al., 2018 | Observational | Statistically significant [Positive (peer)]<br>Statistically significant [Negative (partner)] | Family<br>Partner<br>Peer | During treatment | Methadone | Standard: MMT |
| Wasserman et al., 2000 | Observational | No effect [Positive (family, partner, peer)]<br>No effect [Negative (family, partner, peer)] | Family<br>Partner<br>Peer (non-using and drug-using) | During treatment | Methadone or LAAM | Standard: MMT/LAAMMT |
| Yandoli et al., 2002 | Experimental | Statistically significant | Family<br>Partner | During treatment | Methadone | Supportive psychotherapy + standard treatment<br>Family therapy + standard treatment<br>Standard: MMT (called low contact treatment) |
| Yang et al., 2013 | Observational | Statistically significant [Positive (family, partner)]<br>Statistically significant [Negative (family, partner, peer)] | Family<br>Partner | Pre-treatment | Methadone | Standard: MMT |
| Zhou et al., 2017 (1) | Observational | Statistically significant | Family<br>Partner<br>Peer | Pre-treatment | Methadone | Standard: MMT |

Table 5: Synthesis (continued)
| Study | Study type | Statistical significance of outcome at $p < .05$ | Form of social network support | Point in treatment timeline that social network is assessed | MOUD drug used | Intervention (if any) |
| --- | --- | --- | --- | --- | --- | --- |
| Zhou et al., 2017 (2) | Observational | Statistically significant [Positive (family, partner)]<br>Statistically significant [Negative (peer)] | Family (cohabiting)<br>Partner (cohabiting)<br>Peer (drug-using) | Pre-treatment | Methadone | Standard: MMT |
| Zhu et al., 2018 | Observational | Statistically significant | Family<br>Partner<br>Peer | Post-treatment | Methadone or buprenorphine/naloxone | Standard: randomized to either MMT or buprenorphine/naloxone |

Abbreviations: BMT - Buprenorphine maintenance treatment, CM - Contingency management, LAAM - Levo-alpha-acetylmethadol harm reduction, MMT - Methadone maintenance treatment, RCT - Randomized controlled trial
Note: Observational studies do not have randomized treatment and therefore don't have impact of bias
Note: For the Statistical significance of outcome at $p < .05$ column, we denoted the statistical significance of positive and negative ties if such information was provided (see Table 2)

## Discussion

Despite the vast body of research conducted on the use of medication for opioid use disorder, there are still wide gaps in our understanding of how the social network environments of patients may mediate the success of various treatment approaches. Given the persistent impact of the opioid epidemic, it is critical that those who are exploring ways to improve MOUD access and outcomes––from clinicians to politicians––are informed of the significant role that certain social relationships play in those undergoing treatment in community settings.

This systematic review represents the current body of knowledge on the role of social network support and social network ties on MOUD treatment outcomes. The effect of three types of social network relationships––family, partner, and peer––categorized at three time points––pretreatment, during treatment, and post-treatment––was recorded with respect to MOUD treatment outcomes. Fourty-six observational and experimental studies over a range of social network support variants were considered. Results from the reviewed studies generally support the idea that certain characteristics of social network ties can have positive and negative associations with treatment outcomes, although some non-significant findings indicate that more research is necessary to firmly establish the relationship between network support and MOUD treatment outcomes. Variation in results may also be attributed to the variability of outcome types, differences in comparison groups, and different pharmacological interventions. Due to the variation in outcomes, a meta-analysis of experimentally-driven studies was not possible.

The bulk of the studies considered in this review evaluated pre-treatment or during treatment sources of social support and were relatively consistent with the dynamic network framework proposed in Fig 1. Such a framework is based on patterns in the literature and holds that, at a basic level, a patient’s successful treatment outcome in MOUD will be accompanied by a shift in their network composition away from the drug-using network and towards other relationships that are significant sources of social support throughout treatment. Observational studies relating social relationship factors before and during treatment to outcomes are valuable for two reasons: first, because they reveal patterns in the social network composition of individuals entering MOUD programs, and second, because levels of support at treatment initiation may be predictive of treatment success at a later time point. The picture at these time points provided by the studies reviewed here suggests that, in general, patient social support from non-drug-using network individuals is low at treatment initiation while contact with the drug-using network is high. With considerable evidence that social support, fewer relationship conflicts, and little exposure to other substance users are positively associated with desired treatment outcomes, future efforts may utilize such observational methods to identify individuals at-risk of terminating treatment early on. Overall, there were few studies which followed and documented patient social network support after discharge and its relation to continued abstinence, despite a major challenge to MOUD being long-term recovery, demonstrating an area for further study.

The experimental studies reviewed here actively involved other individuals in the patient’s network in treatment; results suggested that outcomes were generally improved using such interventions. Despite the wide body of work on the efficacy of administering psychosocial interventions in addition to pharmacological treatment, and the numerous reviews to this effect [16, 25, 26], there is very little work looking into psychosocial interventions which specifically leverage significant others in patient networks. While two studies utilized network mapping methods to identify others to participate in therapy [41, 67], the most prominent positive effects of interventions involving network members seemed to be relationships-specific, such as couples therapy between partners [66, 60] or parent skills training between parents and children [76, 72].

While the results of our review did not decisively indicate whether a particular social network relationship was inherently influential, the evidence base for family social network support appears the most significant. It should be noted that the importance of relationships are themselves mediated by other factors, such as local norms. Family social network support was particularly emphasized in East Asian countries, where family assumes a particularly important role in a patient’s life [46, 47, 48, 49, 54]. Future research might explore how certain relationship involvement in treatment can leverage local socio-cultural norms to improve MOUD outcomes. More generally, many of the studies reviewed here reveal that characteristics of relationships, and not the particular types of relationships, may be the most influential aspects of social ties during treatment. For example, evidence from several studies across different time points suggests that, regardless of the type of relationship, having and maintaining social ties during treatment that are non-judgemental of MOUD or abstinence-positive [54, 77, 68, 60, 76, 63], that have little conflict [65, 64, 53], that are mutually non-substance-using [69, 43, 74, 78], or that involve cohabitation [61, 50, 56] is positively associated with successful treatment outcomes.

Given that the majority of the studies reviewed here are consistent with our theoretical framework for how social network changes accompany successful treatment, future work might also aim to further investigate network change over time in the community clinic setting. With networks in mind, the hypothesized role of relationships in treatment is complex, as perceptions of family, partner, and peers may directly impact treatment, may indirectly impact treatment through a mediator such as motivation to attend clinic, may be altered by treatment progression, or there may be a combination of above factors [44]. Nonetheless, past research on social networks of both tobacco and alcohol users has indicated that there is significant contagion of both use and quitting behavior between social contacts [18, 79], warranting further investigation of opioid use and treatment through the social network lens. However, network change approaches may not be realistic if, for example, participants are trying to decrease contact with another substance user that is a close friend or cohabitee. Leveraging findings from this review, it may be possible engage patients with such networks in disseminating harm reduction-centric and abstinence-positive information around opioid use to their social network during treatment. This may extend the reach of social network support interventions in not only improving patient outcomes, but also improving broader community well-being. Interventions might also be selectively geared towards social tie creation for patients who have few social contacts at baseline. One study that encouraged participants who did not have a drug-free contact in their network to attend community activities like church noted the potential positive impact of such engagement in creating new drug-free ties, though the effect on outcomes in the study was non-significant potentially due to low attendance [69].

Our findings are of great relevance given the current gap in understanding of MOUD treatment outcomes, and particularly those problems raised by rates of high attrition and high illicit substance use concurrent with treatment. The results presented here suggest that social network information can be utilized to personalize existing interventions or leveraged to innovate new approaches. With psychosocial interventions that supplement pharmacological treatment becoming the norm in combating opioid dependence, it is critical that researchers develop the most efficacious MOUD interventions. The current review also provides wider policy implications. Governments and public health authorities seeking to improve treatment outcomes might incorporate successful, network-targeting MOUD interventions noted in our review. Public health authorities can encourage treatment providers to explore social network support or other similar techniques to augment MOUD. Such agencies might also incentivize treatment providers to include different forms of social network support in their programs to determine which is most efficacious and simultaneously build upon the research base of network support in MOUD approaches. Similarly, in absence of a large evidence base for social network support-based MOUD approaches, treatment providers with generic approaches (i.e., without any form of behavioral/psychosocial intervention) might informally encourage patients to seek social support from their family, friends or peers.

Most of the studies had key methodological concerns. We emphasized more robust study designs and assessed the probable impact of bias to compensate for methodological weaknesses. Possible sources of bias included group baseline differences, selection bias, attrition bias, and differential rates of follow-up. Selection bias may have exaggerated or under-reported treatment effects. If rates of attrition were relatively high or greater in untreated groups, there may be a possibility that treatment effects were overestimated if participants lost to follow-up had greater negative outcomes. A few studies in this review conducted analysis to control for bias through multivariate analysis and/or comparison of losses to follow-up with those followed-up. Attempts to account for biases may not always be successful, and we thus assessed the risk of biases (see Table 4), providing an assessment of probable impact of bias on various outcomes.

Limitations also arose from differences in methods of reviewed studies, making it more complex to assess or synthesize all studies under the same rubric. The details provided on methods and analysis was highly varied, possibly leading to fluctuations in the confidence level of results. Finally, there were limitations in our classification method that arose from the high variability between study methods and analysis. For example, because assumptions had to be made about relationships assessed in some studies, some results may be based primarily one one relationship despite a distinction not being drawn in the original study and therefore not drawn here (e.g., study describing family and partner support in which vast majority of participants elicit support from partner). Additionally, peer relationships in particular were limited by definition in our review, often unspecified or generally specified as friends. Future studies might aim to assess ‘weak’ peer ties, such as co-workers, employers, or clinic care providers.

## Conclusions

Although evidence was occasionally mixed, social network support and social tie influence measured across various points in the treatment timeline was generally related to MOUD treatment outcomes. Interventions around social network support could potentially augment MOUD treatment outcomes, possibly playing a role in mitigating the current epidemic of opioid dependence and overdose. Future research should explore social network changes across the treatment timeline and leverage patterns in the changes to address obstacles to MOUD success.

## Data Availability

All journals reviewed are publically indexed.

