## Supplementary Information for "The role of social network support in treatment outcomes for medication for opioid use disorder: a systematic review"

#### Search strategy

We searched online indexes, references in previous reviews/guidelines, and Clinicaltrials.gov. In addition, we consulted content experts. We conducted a systematic review of the literature using the databases of PubMed, MEDLINE, Embase, PsycINFO and Sociological Abstracts. We searched literature from inception through August 2020. Studies written in English, conducted with humans, mentioned MOUD in the title or abstract, included social network support were considered for inclusion. Additional studies were identified by scanning reference lists of previous literature reviews and other studies. To reduce publication bias, we included a broad range of studies [1]. The ClinicalTrials.gov library was searched to identify potentially qualifying studies that have not led to published results. We obtained additional papers through consultation with experts and authors, targeted searches of thematic journals, technical reports, conference proceedings and national databases.

The following is an example of our PubMed search strategy:

((”buprenorphine, naloxone drug combination”[MeSH Terms] OR ”naltrexone”[MeSH Terms] OR ”methadone”[MeSH Terms] OR ”opiate substitution treatment”[MeSH Terms] OR ”opioid-related disorders”[MeSH Terms] OR ”medication assisted therapy”[tw] OR ”medication assisted therapies”[tw] OR Naltrexone[tw] OR Methadone[tw] OR Celupan[tw] OR Trexan[tw] OR ReVia[tw] OR Nemexin[tw] OR Nalorex[tw] OR Antaxone[tw] OR ”EN 1639A”[tw] OR Vivitrol[tw] OR Suboxone[tw] OR ”Buprenorphine Naloxone”[tw] OR Methadone[tw] OR Dolophine[tw] OR Metadol[tw] OR Symoron[tw] OR Methadose[tw] OR Phenadone[tw] OR Physeptone[tw] OR Phymet[tw] OR Amidone[tw] OR Methaddict[tw] OR ”Methadone Maintenance Treatment”[tw] OR ”Opiate Substitution Treatments”[tw] OR ”Opioid Substitution Treatment”[tw] OR ”Opioid Substitution Treatments”[tw] OR ”Opioid Substitution Therapy”[tw] OR ”Opioid Substitution Therapies”[tw] OR ”Opiate Replacement Therapy”[tw] OR ”Opiate Replacement Therapies”[tw] OR ”Opioid Replacement Therapy”[tw] OR ”Opioid Replacement Therapies”[tw]) AND (”social support”[MeSH Terms] OR ”community health services”[MeSH Terms] OR ”community networks”[MeSH Terms] OR ”spouses”[MeSH Terms] OR ”friends”[MeSH Terms] OR ”family”[MeSH Terms] OR ”societies”[MeSH Terms] OR ”residence characteristics”[MeSH Terms] OR ”social support”[tw] OR ”social supports”[tw] OR ”social network”[tw] OR ”social networks”[tw] OR ”support system”[tw] OR ”support systems”[tw] OR Spouse[tw] OR Spouses[tw] OR Partner[tw] OR Partners[tw] OR Friend[tw] OR Friends[tw] OR Society[tw] OR Community[tw] OR Communities[tw] OR Peer[tw] OR Peers[tw] OR Family[tw] OR Families[tw] OR

Husband[tw] OR Husbands[tw] OR Wife[tw] OR Wives[tw] OR co-worker[tw] OR co-workers[tw] OR coworker[tw] OR coworkers[tw] OR neighbor[tw] OR neighbors[tw] OR Neighborhood[tw] OR Neighborhoods[tw] OR Neighbourhood[tw] OR neighbourhoods[tw])) NOT ((”buprenorphine, naloxone drug combination”[MeSH Terms] OR ”naltrexone”[MeSH Terms] OR ”methadone”[MeSH Terms] OR ”opiate substitution treatment”[MeSH Terms] OR ”medication assisted therapy”[tw] OR ”medication assisted therapies”[tw] OR Naltrexone[tw] OR Methadone[tw] OR Celupan[tw] OR Trexan[tw] OR ReVia[tw] OR Nemexin[tw] OR Nalorex[tw] OR Antaxone[tw] OR ”EN 1639A”[tw] OR Vivitrol[tw] OR Suboxone[tw] OR ”Buprenorphine Naloxone”[tw] OR Methadone[tw] OR Dolophine[tw] OR Metadol[tw] OR Symoron[tw] OR Methadose[tw] OR Phenadone[tw] OR Physeptone[tw] OR Phymet[tw] OR Amidone[tw] OR Methaddict[tw] OR ”Methadone Maintenance Treatment”[tw] OR ”Opiate Substitution Treatments”[tw] OR ”Opioid Substitution Treatment”[tw] OR ”Opioid Substitution Treatments”[tw] OR ”Opioid Substitution Therapy”[tw] OR ”Opioid Substitution Therapies”[tw] OR ”Opiate Replacement Therapy”[tw] OR ”Opiate Replacement Therapies”[tw] OR ”Opioid Replacement Therapy”[tw] OR ”Opioid Replacement Therapies”[tw]) AND (”social support”[MeSH Terms] OR ”community health services”[MeSH Terms] OR ”community networks”[MeSH Terms] OR ”spouses”[MeSH Terms] OR ”friends”[MeSH Terms] OR ”family”[MeSH Terms] OR ”societies”[MeSH Terms] OR ”residence characteristics”[MeSH Terms] OR ”social support”[tw] OR ”social supports”[tw] OR ”social network”[tw] OR ”social networks”[tw] OR ”support system”[tw] OR ”support systems”[tw] OR Spouse[tw] OR Spouses[tw] OR Partner[tw] OR Partners[tw] OR Friend[tw] OR Friends[tw] OR Society[tw] OR Community[tw] OR Communities[tw] OR Peer[tw] OR Peers[tw] OR Family[tw] OR Families[tw] OR Husband[tw] OR Husbands[tw] OR Wife[tw] OR Wives[tw] OR co-worker[tw] OR co-workers[tw] OR coworker[tw] OR coworkers[tw] OR neighbor[tw] OR neighbors[tw] OR Neighborhood[tw] OR Neighborhoods[tw] OR Neighbourhood[tw] OR neighbourhoods[tw])).
Specialist journals searched included: Addiction, Addiction Research  Theory, Addictive Behaviours, American Journal of Addictions, Addiction Science and Clinical Practice, Drug and Alcohol Dependence, Drug and Alcohol review, Drugs: Education, Prevention and Policy, European Addiction Research, International Journal of Drug Policy, Journal of Addiction Medicine, Journal of Addiction and Offender Counselling, Journal of Alcohol and Drug Education, Journal of Drug Issues, International Journal of Mental Health and Addiction, Addictive Disorders and Their Treatment, Journal of Substance Abuse, Journal of Substance Abuse and Treatment, Journal of Substance Use, Substance Abuse: Research and Treatment, Substance Abuse Treatment, Prevention and Policy and Substance Use  Misuse.

### Data extraction

We utilized a standardized template to extract data from each study. We extracted general information (e.g., year, setting) and methods (e.g., design, duration), variant of MOUD (e.g., methadone, buprenorphine, naltrexone), and results specific to each outcome (e.g., treatment adherence, self-reported drug use, urine drug screen). Endnote, a bibliographic software, was used to store, organize and manage all references [2]. Covidence was used to manage the screening phases [3].

The authors, in groups of two, independently conducted study selection. A standardized template was pre-piloted independently by two authors, and all authors, in groups of two, extracted all relevant data. The authors resolved disagreements in study selection and data extraction through discussion. A third author stepped in when necessary for a final arbitration of any disagreements that occurred. In groups of two, the authors independently evaluated quality assessments and outcomes for each study and reached consensus via discussion. When consensus was not reached, a third reviewer made final decisions. Quality assessments for experimental studies were conducted using criteria from the Cochrane Handbook [4] and similarly described the quality of observational studies.

We assessed possible bias arising from low or differential follow-up rates, as losses to follow-up may have more negative outcomes than included participants. We considered potential bias in self-report data due to social acceptability. For experimental studies, assessment included level of randomization, rates of attrition in the experimental group, use of intention-to-treat analysis and how group-level baseline differences were dealt with. For observational studies, bias assessment centered on group similarity (e.g., matching), selection-bias and baseline differences possibly influencing outcomes (e.g., severity of dependence), and on analyses (e.g., multivariate logistic regression) adjusting for pre-study group differences. We assessed risk of bias at the study level or specific outcome level. We detailed whether biases were likely to exaggerate or under-estimate the reported treatment effect.

In non-randomized studies, systematic bias may occur between different strata of social network support. This was a general problem with observational studies because parsing between causal relationships around social network support and severity of treatment outcomes is complex. Inclusion of data from unpublished studies may reduce risk of publication bias. We used a structured narrative format to synthesize the literature, organized by research question and thematic focus.

### References


[1] Mitchell Ojmarrh, Wilson David B, MacKenzie Doris L. The effectiveness of incarceration-based drug treatment on criminal behavior: A systematic review *Campbell systematic reviews.* 2012;8:i–76.

[2] Analytics Clarivate. Endnote X8 for Windows *Philadelphia, PA: Clarivate Analytics.* 2017.

[3] Innovation Veritas Health. Covidence systematic review software 2017.

[4] Higgins Julian PT, Green Sally. *Cochrane handbook for systematic reviews of interventions*;4. John Wiley & Sons 2011.
